# Longitudinal dynamics of mutant huntingtin and neurofilament light in Huntington’s disease: the prospective HD-CSF study

**DOI:** 10.1101/2020.03.31.20045260

**Authors:** Filipe B. Rodrigues, Lauren M. Byrne, Rosanna Tortelli, Eileanoir B. Johnson, Peter A. Wijeratne, Marzena Arridge, Enrico De Vita, Naghmeh Ghazaleh, Richard Houghton, Hannah Furby, Daniel C. Alexander, Sarah J. Tabrizi, Scott Schobel, Rachael I. Scahill, Amanda Heslegrave, Henrik Zetterberg, Edward J. Wild

## Abstract

The longitudinal dynamics of the most promising biofluid biomarker candidates for Huntington’s disease (HD) – mutant huntingtin (mHTT) and neurofilament light (NfL) – are incompletely defined, but could help understand the natural history of the disease and how these biomarkers might help in therapeutic development and the clinic. In an 80-participant cohort over 24 months, mHTT in cerebrospinal fluid (CSF), and NfL in CSF and blood, had distinct longitudinal trajectories in HD mutation carriers compared with controls. Baseline analyte values predicted clinical disease status and subsequent clinical progression and brain atrophy, better than did the rate of change in analytes. Overall NfL was a stronger monitoring and prognostic biomarker for HD than mHTT. Nonetheless, mHTT possesses prognostic value and is a valuable pharmacodynamic marker for huntingtin-lowering trials.

## Introduction

Despite knowledge of its monogenetic cause, no treatments have been shown to slow neurodegeneration in Huntington’s disease (HD)^1,2^. However, multiple approaches aimed at lowering production of the causative mutant huntingtin protein (mHTT) are in human clinical trials^3–5^. The ultimate goal – treating mutation carriers early, to prevent disease onset – will require prevention trials in premanifest HD mutation carriers (preHD).

Successful target engagement by the first targeted huntingtin-lowering therapeutic tested in HD patients – the antisense oligonucleotide tominersen (formerly IONIS-HTTRx/RG6042) – was demonstrated by dose-dependent mHTT reduction in cerebrospinal fluid (CSF) in a phase 1/2 trial^3^, quantified by ultra-sensitive immunoassay^6^. A reliable CSF to brain mHTT relationship has been established in animal studies^7,8^. This notable success led to the first phase 3 trial of such a drug, whose primary outcomes are the Total Functional Capacity (TFC) score of the Unified Huntington’s Disease Rating Scale (UHDRS) in the USA, and a composite UHDRS (cUHDRS) measure combining motor, functional, and cognitive scores in the EU^9^. Such clinical rating scales quantify overt clinical manifestations, but are less sensitive to detect deterioration, or its therapeutic benefit, in preHD^10–13^ making their use as outcomes in prevention trials problematic. Though clinically relevant, they are also far removed from the core disease mechanism: neuronal injury by the *HTT* gene product. Quantifying biochemical manifestations of neurodegeneration can inform our understanding of pathobiology and the development and testing of novel therapies.

To this end, we recently showed that CSF levels of mHTT – the toxic pathogenic protein – and neurofilament light (NfL) – an axonal protein indicative of neuronal injury – are among the earliest detectable changes in HD, and are strongly associated cross-sectionally with baseline measures of clinical severity and brain volume^14^. In the longitudinal Track-HD cohort, we showed that blood NfL level independently predicts subsequent onset, clinical progression, and brain atrophy in HD over three years^15^.

One unexpected finding from the phase 1/2 ASO trial was a transient elevation of NfL in CSF around five months after first dose^3^. In the subsequent open-label extension of this trial^16^, NfL levels rose again then fell in the context of ongoing huntingtin suppression, almost to baseline by month 9^8^. While it remains unexplained mechanistically, this observation attests to the combined value of mHTT and NfL to highlight changes of note in the “undiscovered country” of huntingtin-lowering. It also calls for a more detailed understanding of how these markers change over time throughout the life of mutation carriers.

Here we present the mHTT and NfL findings from the two-year prospective longitudinal HD-CSF study, in which an 80-participant cohort of HD mutation carriers and controls underwent clinical assessments, sampling of CSF and plasma, and MR imaging, under strictly standardised conditions. To our knowledge, this is the first report of the longitudinal dynamics of CSF mHTT and NfL, studied and compared head-to-head in the natural history of HD. We assessed and compared the ability of the biomarkers at baseline, and their rates of change, to predict longitudinal progression in clinical and neuroimaging measures. We employed advanced methods including random forest methodology and event-based modelling, permitting the exploration of non-linear relationships and multiple variables at once. Finally, we performed computational clinical trial simulations to provide insight into how they could be used and combined in the therapeutic context.

## Results

### The HD-CSF Cohort

Seventy-four (92.5%) out of the eighty baseline participants returned for the 24-month follow-up assessments. Three (4%) out of the seventy-four opted out of doing the follow-up lumbar puncture but agreed to blood and phenotypic data collection (Figure 1). A more detailed version of the study flow is provided in Supplementary Fig. 1.

**Figure 1.**
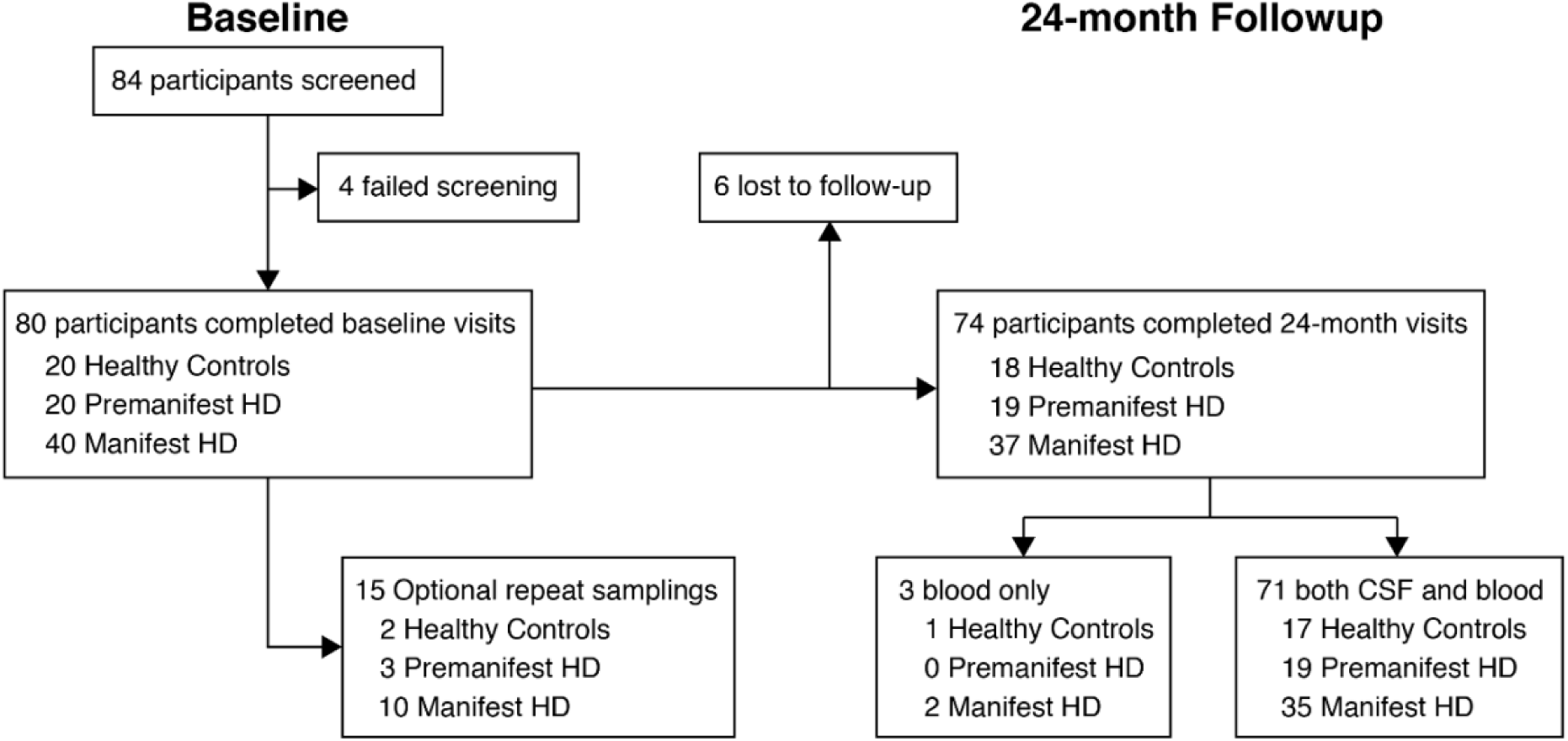
HD-CSF study participant disposition. Baseline visit (n=80) was performed 24-months (± 3 months) before the follow-up visit (n=74). Optional repeat sampling visits occurred 6-8 weeks after baseline. A more detailed version including all study assessments is provided in Supplementary Fig. 1.

Full cohort characteristics are presented in Supplementary Table 1. Disease groups were well-matched for gender and differed as expected in HD clinical, cognitive and imaging measures. Age differed significantly between groups due to the control group (50.68 years ± 11.0) being matched to all HD mutation carriers, and manifest HD (56.02 years ± 9.36) being more advanced in their disease course than preHD (42.38 years ± 11.04), as previously reported^14^.

### Longitudinal dynamics of mHTT and NfL

Longitudinal trajectories of each analyte within individuals are shown in Figure 2a-c. For NfL in CSF and plasma, there was little overlap in the trajectories of HD mutation carriers and healthy controls. Mixed-effects models depict distinct patterns of longitudinal analyte dynamics (Figure 2d-f). CSF and plasma NfL showed a sigmoidal pattern over time in HD mutation carriers, initially accelerating then later slowing, compared to a slow linear rise with ageing in controls. CSF mHTT, on the other hand, rose linearly with age, albeit with more variability.

**Figure 2.**
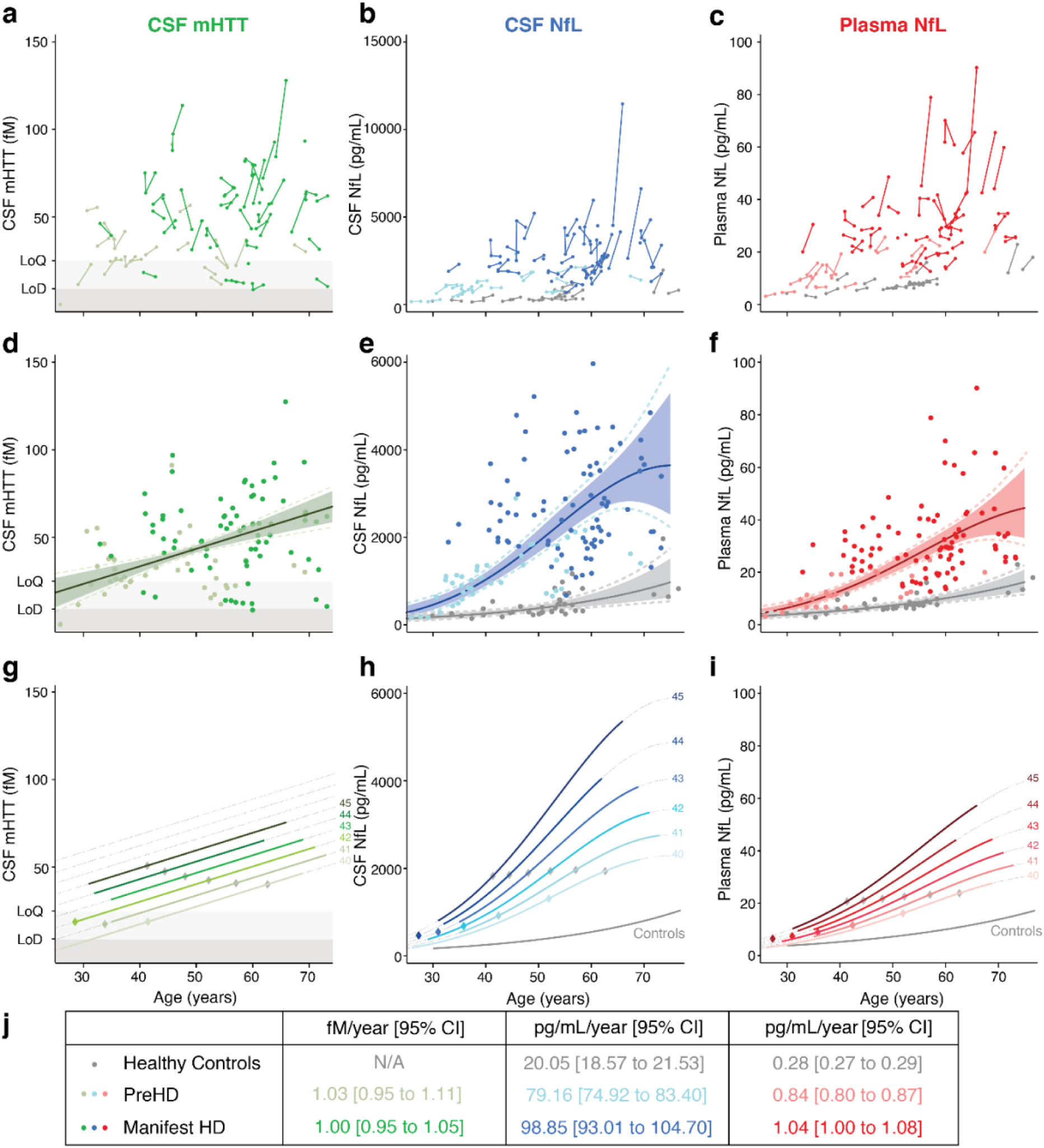
The longitudinal dynamics of mHTT and NfL over 24 months. (a-c) Individual participant trajectories. Connected dots are measurements within the same participant. Disease groups are colour coded as per (j). (d-f) Modelled biomarker trajectories. Model (solid line), 95% (coloured area) and 99% (dashed line) bias-corrected accelerated confidence intervals were generated from generalized mixed effects models. For CSF mHTT, age was used as first-order fixed effect, while for HD mutation carriers for NfL there was a first- and second-order fixed effect for age. All models were adjusted for CAG repeat count and had a random intercept for participant, and corresponding random slopes for age. Dots represent the observed values. For ease of visual interpretation 2 individual data points (>6,000 pg/mL) were included in the model but excluded from figure (e). (g-i) Modelling genetic dose-response relationships to show associations between biomarkers, age and CAG repeat count. For all the analytes, the linear combinations of the interactions between age factors and CAG repeat were significant (CSF mHTT p=0.002; CSF NfL p=0.008; plasma NfL p=0.001). Solid lines were produced from our observations using the models above; dashed lines are predictions outside the range of our observations. Separate figures with individual data points for each individual CAG repeat count are provided in Supplementary Fig. 2. Grey diamonds show the age of predicted onset for each CAG length (as per Langbehn et al. 200417). Coloured diamonds show the age at which HD mutation carrier trajectories are most likely to depart from healthy controls trajectories for each CAG repeat count, generated by change-point analysis. (j) Annualised rates of change and 95% confidence intervals. For each biomarker, estimates were computed as the average of the rate of change in 1,000 simulations per group of study participants (i.e. healthy controls, premanifest and manifest HD). For CSF mHTT, shaded areas mark the limits of detection (LoD, 8 fM) and quantification (LoQ, 25 fM) of the assay. CSF, cerebrospinal fluid; mHTT, mutant huntingtin; N/A, not applicable; NfL, neurofilament light.

To explore the ‘genetic dose-response relationship’ between the causative gene mutation and each biofluid measure we queried our models for the interaction between CAG and age in HD mutation carriers (linear effect for CSF mHTT, p<0.0001; and nonlinear effect for CSF and plasma NfL, p=0.006 and p<0.001, respectively; Figure 2g-i; Supplementary Fig. 2). Using a change-point analysis, we estimated the disease burden score (a combination of age and CAG) at which each analyte in HD mutation carriers starts to deviate from controls. The DBS change-points for each analyte (CSF mHTT, 188.9; CSF NfL, 248.6; and Plasma NfL, 236.1; Supplementary Fig. 3) were used to annotate Figure 2g-i with the age of expected departure from controls, for each CAG length.

Based on simulations of the models in Figure 2d-f, CSF NfL rose fastest in manifest HD by 98.85 pg/mL/year, in preHD by 79.16 pg/mL/year and in controls by 20.05 pg/mL/year (Figure 2j). Similar relative findings were seen for plasma NfL rates of change (controls 0.28 pg/mL/year, preHD 0.84 pg/mL/year, manifest HD 1.04 pg/mL/year; Figure 2j).

### Prognostic value for overall HD progression of baseline analyte versus its rate of change

We assessed the clinical associations of each analyte using the cUHDRS, a composite score derived from large natural history cohorts and combining motor, functional and cognitive symptoms to reflect overall HD clinical severity across clinically important domains^18^. All three analytes had significant associations with cUHDRS cross-sectionally at both baseline and follow-up (Extended Data Fig. 1). To assess the prognostic value of each analyte for HD progression, we first examined whether their baseline values predicted subsequent change in cUHDRS. Significant associations with subsequent cUHDRS change were found for all three (CSF mHTT r=-0.31, 95%CI -0.57 to -0.03, p=0.026; CSF NfL r=-0.38, 95%CI -0.52 to -0.18, p<0.0001; plasma NfL r=-0.47, 95%CI -0.63 to -0.25, p<0.0001; Figure 3a-c). The association with baseline plasma NfL remained significant after adjustment for age and CAG (CSF mHTT r=-0.11, 95%CI -0.48 to 0.18, p=0.513; CSF NfL r=-0.21, 95%CI -0.48 to 0.00, p=0.098; plasma NfL r=-0.33, 95%CI -0.58 to -0.08, p=0.011).

**Figure 3.**
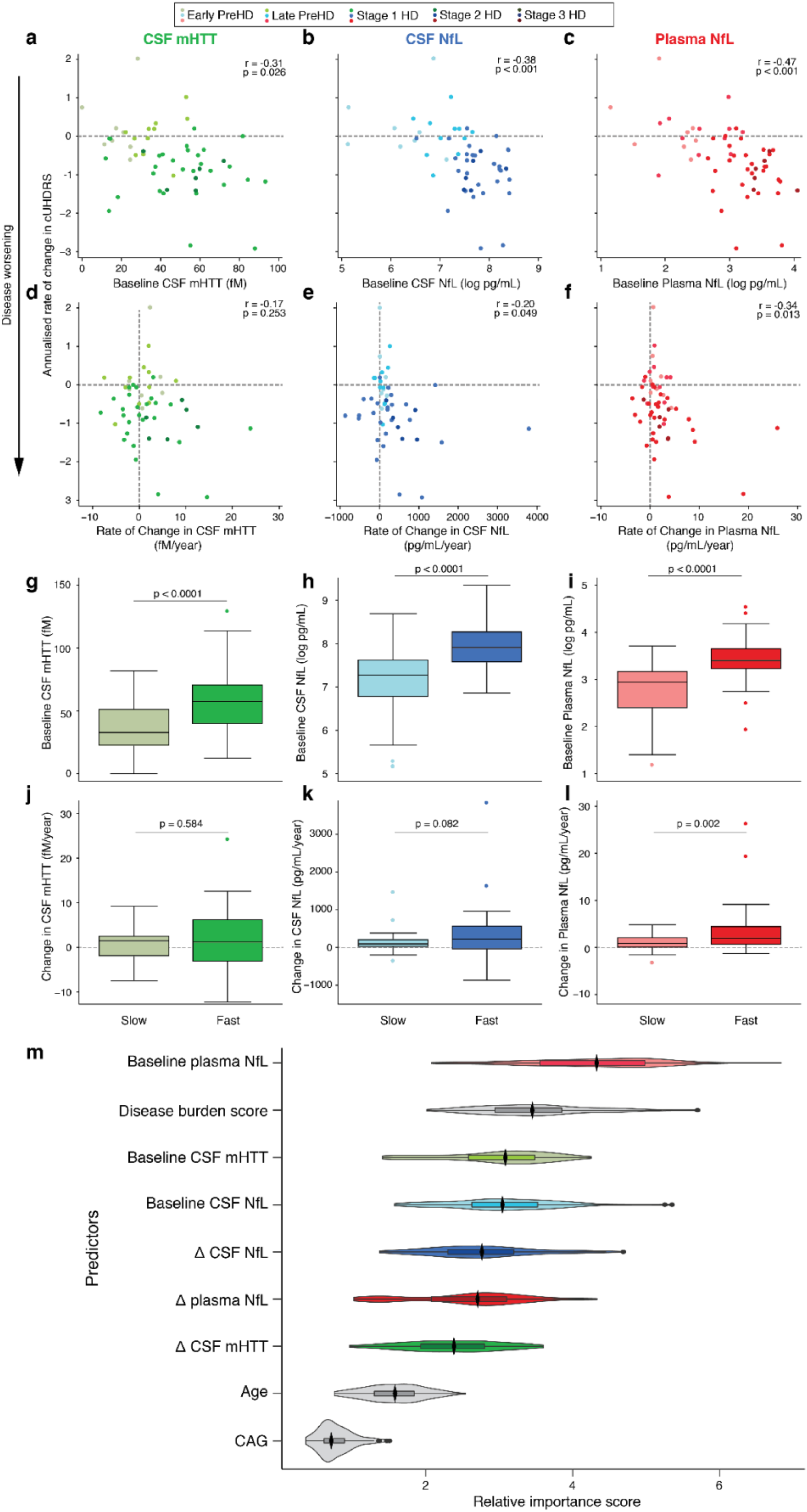
Longitudinal associations of mHTT and NfL with disease progression quantified by cUHDRS. Associations between (a-c) baseline values or (d-f) annualised rate of change in each analyte and the annualised rate of change in the cUHDRS. Partial Pearson’s correlation coefficient adjusted for age, and p-values are presented. Dashed horizontal lines mark no change in cUHDRS, with negative values representing deterioration. Dashed vertical lines mark no change in the biomarker. The baseline values (g-h) and annualised rate of change (i-l) for each biomarker compared between ‘fast’ (n=24) and ‘slow’ progressors (n=30), defined as participants with an absolute decrease in cUHDRS greater than or equal to 1.2 over the follow-up period. (m) Random forest plot for prediction of annualised rate of change in cUHDRS including baseline and rate of change of biofluid biomarkers as predictors. Higher relative importance score indicates a greater relative importance of the variable in predicting worsening cUHDRS score. Relative importance scores were based on mean decrease in Gini score. Distributions were generated from re-running the model 100 times. The boxes show the median, and 25% and 75% percentiles, while whiskers are the lower and upper adjacent values (i.e. 1.5 times the interquartile range minus the 25% percentile or plus the 75% percentile). Dots are values under and above the adjacent values. Violin plots represent ranking distributions. Black diamonds represent the median. CSF, cerebrospinal fluid; cUHDRS, composite Unified Huntington’s Disease Rating Scale; mHTT, mutant huntingtin; NfL, neurofilament light; DBS, disease burden score; Δ, annualised rate of change.

Next, we assessed whether the rate of change of each analyte gave any additional prognostic information beyond that given by a single baseline measurement. The rate of change in plasma NfL was associated with the rate of change in cUHDRS whereas CSF NfL and mHTT showed weaker associations (CSF mHTT r=-0.17, 95%CI -0.49 to 0.08, p=0.253; CSF NfL r=-0.20, 95%CI -0.43 to -0.01, p=0.049; plasma NfL r= -0.34, 95%CI -0.65 to -0.12, p=0.013; Figure 3d-f). These associations for rate of change were weaker than those of the baseline values, and did not survive adjustment for age and CAG (CSF mHTT r=-0.09, 95%CI -0.46 to 0.15, p=0.539; CSF NfL r=-0.09, 95% CI -0.33 to 0.10, p= 0.401; plasma NfL r=-0.22, 95%CI -0.54 to -0.00, p=0.111).

In our analysis of fast and slow progressors (≥ or < 1.2 decline in cUHDRS respectively), baseline CSF mHTT, CSF NfL and plasma NfL were all significantly higher in faster progressors (mean differences: CSF mHTT 19.27 fM, 95%CI 10.74 to 27.80, p<0.0001; CSF NfL 1066.05 pg/mL, 95%CI 0. 532.62 to 1599.48, p<0.0001; plasma NfL 11.44 pg/mL 95%CI 6.45 to 16.43, p<0.0001; Figure 3g-i). Only plasma NfL remained associated after adjustment for age and CAG (mean differences: CSF mHTT 7.38 fM, 95%CI -0.92 to 15.68, p=0.081; CSF NfL 449.75 pg/mL, 95%CI -61.52 to 961.02, p=0.087; plasma NfL 5.59 pg/mL, 95%CI 1.24 to 9.93, p=0.032).

We repeated this analysis using the rate of change in each analyte. Only the rate of change of plasma NfL was significantly higher in faster progressors (mean differences: CSF mHTT 0.67 fM/year, 95%CI -1.75 to 3.09, p=0.584; CSF NfL 230.23 pg/mL/year, 95%CI -29.32 to 489.78, p=0.082; plasma NfL 2.83 pg/mL/year, 95%CI 1.03 to 4.62, p=0.002; Figure 3j-l). This did not survive age and CAG adjustment (mean differences: CSF mHTT -1.04 fM/year, 95%CI -3.69 to 1.61, p=0.440; CSF NfL 27.07 pg/mL/year, 95%CI -253.56 to 307.70, p=0.849; plasma NfL 1.22 pg/mL/year 95%CI -0.71 to 3.15, p=0.210).

We next used random forest analysis to compare head-to-head and illustrate the relative importance ranking of the three biofluid analytes – including their baseline value and annualised rate of change – in prediction of HD clinical progression, alongside other established predictors (age, CAG and DBS). Using change in cUHDRS as a continuous outcome, baseline values ranked as stronger predictors than annualised rates of change (Figure 3m). Similar results were obtained for prediction of fast versus slow progression (Supplementary Fig. 5a).

### Prognostic value of biofluid clinical, imaging and cognitive measures

Next, we compared the prognostic power of each analyte, in terms of both its baseline value and its rate of change, to predict progression in individual clinical and MRI measures (Figure 4obj; Supplementary Fig. 5; Supplementary Table 2a). Baseline measurements of all analytes had significant associations with subsequent decline in clinical and imaging measures (Figure 4a; Supplementary Fig. 5). Baseline CSF mHTT was associated with worsening in TFC (r=0.40, p=0.001) and atrophy of whole-brain (r=0.38, p=0.008), white-matter (r=0.64, p<0.0001), grey-matter (r=0.59, p<0.0001) and caudate (r=0.45, p=0.002), but had very weak associations with TMS and cognitive measures (r<0.20, p>0.242). Baseline NfL, in both CSF and plasma, was associated with progression in all measures except TMS (CSF NfL: TFC r=0.39, p<0.0001; SDMT r=0.20, p=0.039 ; SWR r=0.22, SCN r=0.26, p=0.016; VFC r=0.23, whole-brain r=0.44, p<0.0001; white-matter r=0.58, p<0.0001; grey-matter r=0.37, p=0.002; caudate r=0.47, p<0.0001; Plasma NfL: TFC r=0.46, p<0.0001; SDMT r=0.32, p=0.009; SCN r=0.30, p=0.014; whole-brain r=0.56, p<0.0001; white-matter r=0.63, p<0.0001; grey-matter r=0.47, p<0.0001; caudate r=0.42, p<0.0001). After age and CAG adjustment (Supplementary Fig. 6; Supplementary Table 2b), associations remained significant between baseline CSF mHTT and subsequent change in white-matter (r=0.45 p<0.0001), grey-matter (r=0.46 p=0.0001) and caudate (r=0.29 p=0.074); between baseline CSF NfL and subsequent change in TFC (r=0.25, p=0.032), white-matter (r=0.37, p=0.009) and caudate (r=0.32, p=0.007); and between baseline plasma NfL and subsequent change in TFC (r=0.34, p=0.06), SDMT (r=0.29, p=0.023), whole-brain (r=0.45, p<0.0001), white-matter (r=0.46, p<0.0001) and grey-matter (r=0.30, p=0.023).

**Figure 4.**
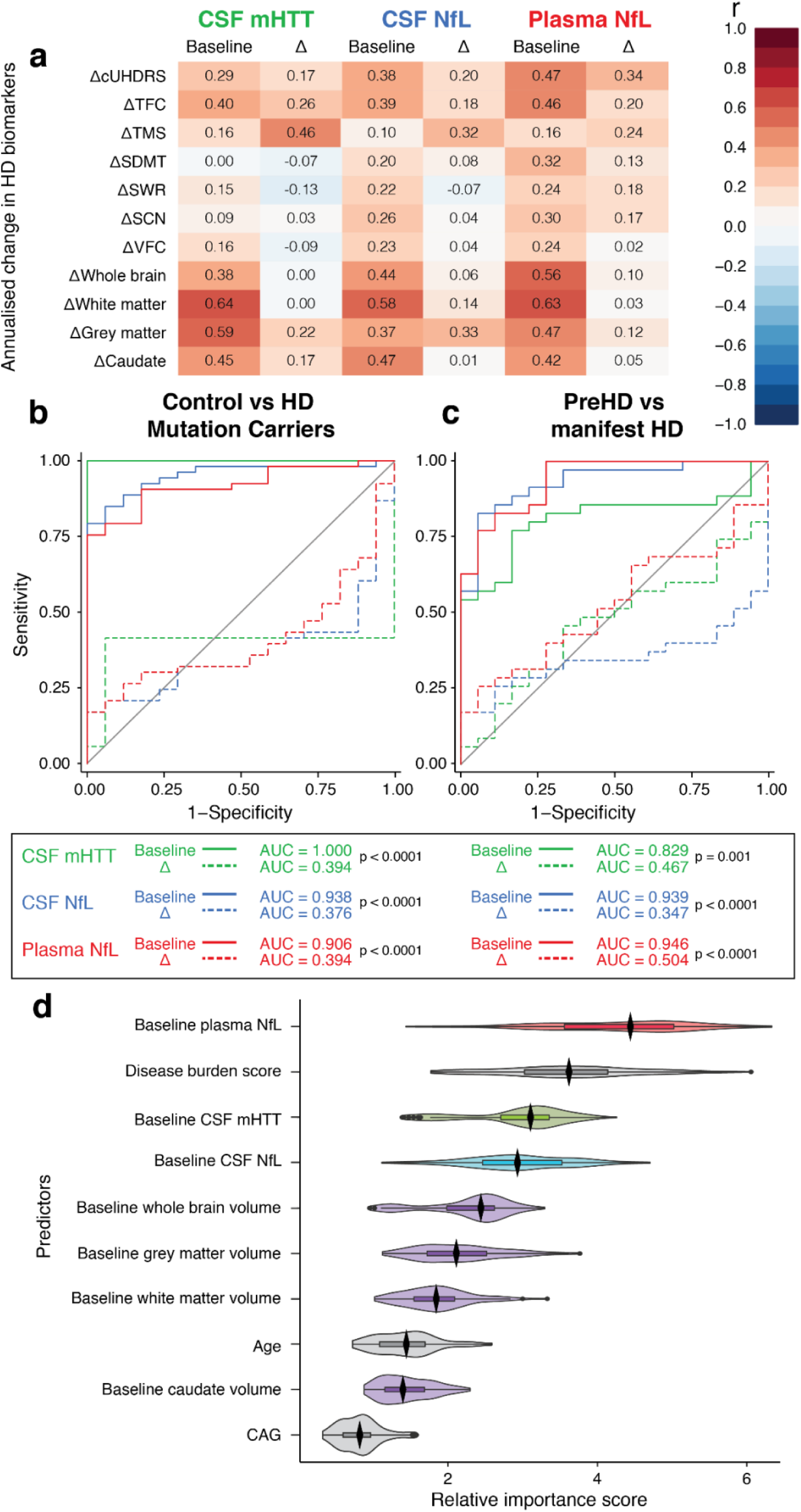
Comparison of prognostic abilities of mHTT and NfL for clinical and imaging measures, and disease state. (a) Matrices show the Pearson’s partial correlation coefficients adjusted for age only for associations between baseline values or annualised rate of change (Δ) of each analyte and the annualised rate of change in clinical and imaging measures, each expressed such that higher positive values denote clinical worsening. Colour coding displays the magnitude and the direction of the association. Coefficients with corresponding confidence intervals and p-values for each combination are provided in Supplementary Table 2. (b-c) ROC curves comparing the discriminatory ability of baseline values for each analyte and its annualised rate of change to distinguish (b) between healthy controls and HD mutation carriers and (c) between premanifest and manifest HD. (d) Random forest plot for prediction of annualised rate of change in cUHDRS including baseline biofluid and imaging biomarkers as predictors. Higher relative importance score indicates a greater relative importance of the variable in predicting worsening cUHDRS score. Relative importance scores were based on mean decrease in Gini score. Distributions were generated from re-running the model 100 times. p-values are for comparison between the baseline AUC and rate of change AUC. Violin plots represent ranking distributions. AUC, area under the curve; CSF, cerebrospinal fluid; cUHDRS, composite Unified Huntington’s Disease Rating Scale; mHTT, mutant huntingtin; NfL, neurofilament light; r, Pearson’s partial correlation coefficient; SDMT, Symbol Digit Modalities Test; SCN, Stroop Color Naming; SWR, Stroop Word Reading; TFC, UHDRS Total Functional Capacity; TMS, UHDRS Total Motor Score; VFC, Verbal Fluency - Categorical; Δ, annualised rate of change.

In contrast, the rate of change in each analyte had weaker associations with progression in every measure apart from change in TMS (mHTT r=0.46, p<0.0001; CSF NfL r=0.32, p=0.012). These associations remained after adjustment for age and CAG (r=0.43, p=0.001; r=0.18, p=0.032 respectively).

Using a receiver operating characteristics (ROC) curve analysis, we compared the discriminatory ability of each analyte’s baseline value and rate of change, to distinguish between different clinical states: controls versus HD mutation carriers, and between premanifest versus manifest HD. For all three analytes, rate of change had poor ability to distinguish in either comparison; areas under the curves (AUCs) were approximately 0.5 (*i*.*e*., no better than chance). Baseline concentrations had excellent discriminatory ability with AUCs greater than 0.8 (Figure 4b,). In each condition, and for each analyte, the AUC for the baseline measurement was significantly greater than that for its rate of change (Figure 4b,c).

To compare the relative prognostic ability of the biofluid and imaging biomarkers within a single model, we repeated the random forest analysis, including only the baseline values for biofluid analytes and imaging biomarkers. Using change in cUHDRS as a continuous outcome, biofluid analytes ranked as stronger predictors than imaging biomarkers (Figure 4d). Similar results were obtained for prediction of fast and slow progression (Supplementary Fig. 4b).

### Simulating clinical trials with biofluid biomarker surrogate endpoints

The data thus far suggests that these analytes indicate current clinical state and have prognostic value for clinical decline. We used longitudinal data from the HD-CSF cohort to run computationally simulated clinical trials using CSF mHTT, CSF NfL and plasma NfL as possible surrogates for clinical progression. These simulations assume that the intervention-induced change in the analyte emulate the change expected in clinical state by an intervention. Figure 5 depicts the relationships between statistical power, sample size and trial duration for such trials, using a nominal 20% drug effect per year on the biomarker trajectory. For longitudinal change in NfL, in CSF or plasma, fewer than 100 participants per arm are needed to show an effect over 9 months. More than 10,000 participants per arm would be required to achieve 80% power for a similar trial using the lowering of CSF mHTT as a surrogate outcome over 24 months. Note that this calculation is based on mHTT release and does not apply to any trial in which the intervention reduces its production directly and lowers the protein below baseline values. Comparable simulations for TFC and cUHDRS, commonly used as clinical trial endpoints, are shown in Extended Data Fig. 2.

**Figure 5.**
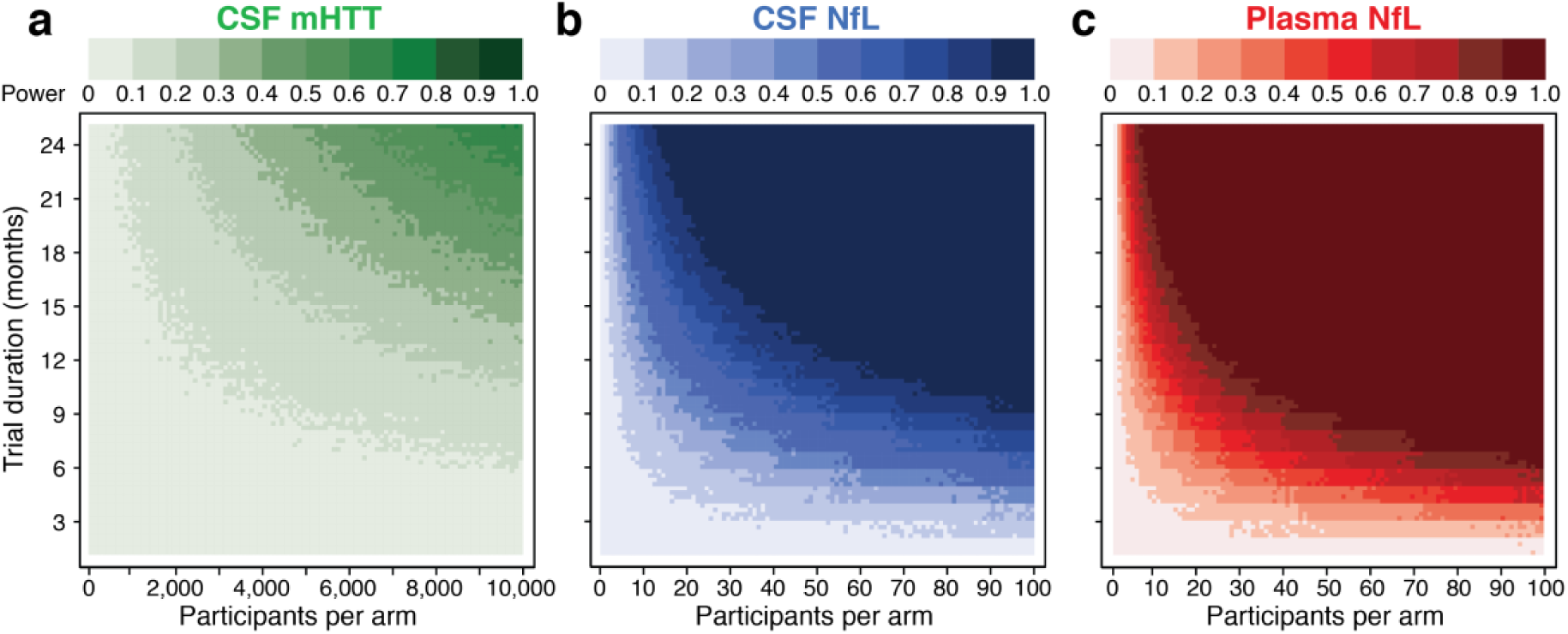
Statistical power, sample size and trial duration. Monte Carlo simulations predicting the statistical power of each biomarker in the clinical trial context, contingent on sample size per arm and trial duration in months. 2-arm parallel design clinical trials with no attrition or placebo effect with a constant effect size of 20% reduction in each analyte per year were simulated. Each pixel represents 1,000 simulated clinical trials, generated using generalised mixed effects models shaped to estimate the longitudinal trajectories of each biomarker (as in Figure 2). The main effect in each simulation repetition was calculated as the inter-arm mean difference in the mean change from baseline, using generalised linear models adjusted for CAG. Statistical power was calculated as the proportion of trial simulations with a p-value < 0.05 for the main effect. CSF; cerebrospinal fluid; mHTT; mutant huntingtin; NfL; neurofilament light.

### Technical validation of cross-sectional baseline results across assays

CSF mHTT, CSF NfL and plasma NfL were re-measured in baseline samples using the same methods used for the follow-up samples, in order to perform the longitudinal analysis. Comparing the re-measured values with those previously published at baseline, batch, assay or storage effects did not affect our samples (Supplementary text; Supplementary Fig. 7). We used the re-measured data to replicate our previously published cross-sectional findings^14^. This included each analytes’ inter-group differences (Extended Data Fig. 3); associations with clinical and imaging measures (Extended Data Fig. 4-5); ROC curves and AUC analysis and event-based modelling (EBM; Extended Data Fig. 6). All results for NfL were similar to those previously published^14^. For CSF mHTT, the re-measured values had stronger associations with clinical measures and, where it previously lacked statistically significant association with brain volumes, the re-measured mHTT was now associated with grey-matter and caudate volume (Extended Data Fig. 5; Supplementary Table 3).

### Replication of cross-sectional results in follow-up data

We used the samples and data from the 24-month follow-up to examine whether our cross-sectional findings held true in the same cohort two years later (Extended Data Fig. 7-10). All results for NfL were similar to those previously published and to the re-measured baseline data. For CSF mHTT, we replicated the stronger associations with clinical measures and found further stronger associations with all brain volumes, similar to those for NfL (Extended Data Fig. 8,9; Supplementary Table 4). Our data-driven event-based model for staging participants based on the totality of their baseline data was validated longitudinally, showing that at follow-up nearly all participants had an EBM stage greater than or equal to the baseline EBM (indicating progression; Supplementary Fig. 8).

## Discussion

Here, we present the 24-month results of HD-CSF, a longitudinal study of biofluid biomarkers in HD mutation carriers and matched controls, with longitudinal clinical and MRI data. We have characterised and compared the longitudinal dynamics of mHTT and NfL in CSF in HD for the first time, defining the trajectories of these biofluid biomarkers and the inflection points at which they depart from healthy controls in the natural history of HD. While rates of change in the analytes had some prognostic value, a single measurement at baseline of each analyte exhibited stronger ability to predict subsequent clinical decline, brain atrophy and disease state. How our novel prognostic findings add to what was known about these biomarkers is summarised in context in Table 1. Using clinical trial simulations, we showed that NfL could be used as an outcome measure of neuronal protection and disease progression, to run trials of feasible duration.

**Table 1.** Summary of what this study adds to our previous understanding of the performance of mHTT and NfL as biomarkers for HD. Citations refer to the first work in which each finding was reported; entries in bold are those reported in the present work. HD, Huntington’s disease; C, healthy controls;

| Cross-sectional data | CSF mHTT | CSF NfL | Plasma NfL |
| --- | --- | --- | --- |
| **Higher in HD v C** | Yes<sup>6</sup> | Yes<sup>25</sup> | Yes<sup>15</sup> |
| **Rises with disease stage** | Yes<sup>6</sup> | Yes<sup>26</sup> | Yes<sup>15</sup> |
| **Baseline level associated with clinical severity** | Yes<sup>6</sup> | Yes<sup>14</sup> | Yes<sup>15</sup> |
| **Baseline level associated with brain volume** | **Yes** | Yes<sup>14</sup> | Yes<sup>15</sup> |
| **Longitudinal data** |  |  |  |
| **Baseline level predicts onset** | ? | ? | Yes<sup>15</sup> |
| **Baseline level predicts clinical progression** | **Yes** | **Yes** | Yes<sup>15</sup> |
| **Baseline level predicts brain atrophy** | **Yes** | **Yes** | Yes<sup>15</sup> |
| **Change predicts clinical progression** | **Yes**(TMS only) | **Yes** | **Yes** |
| **Change predicts clinical atrophy** | **Yes** | **Yes** | **Yes** |

mHTT in CSF, and NfL in CSF and plasma, all rose detectably within participants over 2 years. Over the whole course of the disease, mHTT increases rose linearly with age, whereas NfL rose in a more sigmoidal pattern. The disease-associated rise in NfL was more consistent while mHTT was more variable within individuals. The NfL trajectory was distinct from that in healthy controls, with little overlap. This suggests that monitoring these biomarkers against an age-relevant reference range derived from the healthy population could be clinically meaningful. The dynamics of both CSF mHTT and NfL were also CAG-dependent, revealing longitudinally the genetic dose-response relationships that we demonstrated previously for plasma NfL in the TRACK-HD cohort^15^. Change-point analysis identified the approximate age and analyte concentration at which HD mutation carriers became detectably different from controls, for a given CAG repeat length. Defining these points of deflection from the trajectories in healthy controls may help us move towards models based on CAG repeat length that could be used to enrich or stratify clinical trial participants and could eventually be used to personalise treatment approaches^19^.

For the first time to our knowledge, we assessed the clinical prognostic potential of biofluid biomarkers against the cUHDRS – a composite measure derived from large cohort datasets to have high signal-to-noise ratio as a longitudinal measure of disease progression^18^. mHTT and NfL concentrations predicted change in cUHDRS, affirming their potential as biomarkers of HD progression. We show that both CSF mHTT and NfL each possess prognostic value for subsequent clinical decline and brain atrophy, as seen previously for plasma NfL in the TRACK-HD cohort.

For rate of change in analytes, there was an association with change in TMS but no other measures. That the rate of change in each analyte had lesser prognostic and discriminatory power than their baseline values may appear surprising. Longitudinal studies of NfL in other genetic neurodegenerative diseases, including dominantly inherited Alzheimer’s disease^20,21^ (AD) and frontotemporal dementia^22^ (FTD), have revealed that the rate of change in NfL was a stronger predictor of disease progression, with accelerated rate of change in those who converted from presymptomatic to symptomatic. The HD-CSF study was not designed to assess predictors of conversion from premanifest to manifest HD, but the rate of change in plasma NfL did show significant prognostic value in our comparison of fast versus slow progressors. NfL is an axonal protein but not specific to neuronal sub-populations or to a given disease pathology. It is likely that each disease exhibiting neuronal dysfunction will have a distinct longitudinal NfL profile. It is notable that HD has some of the highest elevated levels of CSF NfL compared to other neurological diseases studied to date, greater than levels in AD and FTD which have more rapid clinical progression^23^. A baseline measure encompasses the totality of a disease’s effects up to the point in a person’s life; by comparison, in a slowly progressive disease, even a 2-year change value captures a relatively small further difference.

The relative importance rankings generated from unbiased random forest analyses suggest that a single measurement of plasma NfL has equivalent, if not superior, prognostic value to that of disease burden score – one of the strongest predictors of HD progression. Further, the baseline biofluid biomarkers were stronger predictors of clinical progression than were brain volumes. This is supportive of a single measurement of biofluid biomarkers being closely indicative of the dynamic pathological processes driving disease progression.

Our computational clinical trial simulations offer a novel means to plan future clinical trials that would use the lowering of these biomarkers as surrogate endpoints. They suggest that a trial using plasma NfL with as few as 100 participants per arm run over six months would have over 90% power to show 20% slowing of the expected longitudinal trajectory of the analyte and, under our assumptions, the slowing of clinical decline. Using CSF NfL, the same trial would need to be run over nine months to achieve the same power. This is striking when compared to the several hundred participants per arm required to achieve the same effect size over 24 months for both cUHDRS and TFC – current clinical end points of the ongoing phase 3 huntingtin-lowering trial^9^. A caveat here is that we assume that the effect size lowering in NfL would be equivalent to the same effect size for improvement in clinical outcome. Until there is a clinically efficacious intervention for HD, this hypothesis cannot be tested. Because of a slower rate of increase, and more importantly, the intra-and inter-subject variability in the change over time, in our simulations a much larger participant numbers would be needed to show a deflection in the trajectory of mHTT as a surrogate outcome (not a pharmacodynamic outcome) by slowing disease progression alone. However, given the existing associations of CSF mHTT as a prognostic and pharmacodynamics response marker, it is likely that this measure too will turn out to have some predictive utility in the drug trial context. NfL is less variable and appears to be a better marker of HD progression and prognosis than mHTT. However, it is important to reiterate that mHTT retains its intrinsic value as a direct measure of the causative neurotoxin and as a means of assessing the on-target effects of huntingtin-lowering agents. To achieve this purpose, very small participant numbers are required, as shown by our previous cross-sectional power calculations and findings from the first human trial of such an agent^3,14^. Our novel finding that lower mHTT concentrations predict more slower progression is potentially important in this respect. Importantly, we also show that long-term freezer storage of samples does not adversely impact quantification of mHTT and NfL.

In our previously published baseline analysis^14^, CSF mHTT concentration was not cross-sectionally associated with any brain volume measure. However, in our replication of cross-sectional analyses we found associations between CSF mHTT and caudate and grey-matter volume at baseline, and strong associations with all brain volumes with the samples from the 24-month collection. This was likely because the performance of the mHTT assay, which has been enhanced through its use in clinical trial programs, has improved so as to reduce variability in the latest analysis. Further, baseline CSF mHTT values also predicted subsequent brain atrophy, confirming that mHTT level in CSF *per se* has prognostic potential in HD, beyond its intrinsic appeal as a therapeutic target. It remains important to note that several participants’ mHTT values were below the lower level of quantification of the assay, an indication that more sensitive HTT assays will be needed, especially in the realm of preHD and prevention trials.

Despite being, to our knowledge, the largest longitudinal natural history cohort CSF collection with matched MRI data in HD, the sample number remains modest. We lack the granularity to make predictions of clinical outcome at the individual level. Larger sample numbers and greater follow-up durations may bring us closer to developing models that could inform clinical prognosis. We are also limited to two longitudinal time points. The lack of associations between rate of change in analyte and the rate of clinical decline was likely driven by both sets of rates being derived from the same time period. Having additional time points would permit the comparison of rates of change in analyte with rates of clinical decline in the subsequent time period. For instance, change in NfL in year 1 may predict brain atrophy in year 2. The range of HD mutation carriers in this study does not cover the whole spectrum of HD. This study was not designed to detect the very earliest disease-related alterations, nor what happens in the later stages of disease. Nevertheless, our range is broader than what is currently recruited for clinical trials. Finally, the sample sizes for TFC and cUHDRS generated from the clinical trial simulations were larger compared to ongoing trials. It is important to note that given the range in HD clinical severity, the variability is larger in our cohort, and we used a conservative effect size, all of which drive larger sample size.

Efforts are well underway to address these issues: HDClarity, a multi-national CSF collection initiative for HD has amassed over 600 CSF and plasma samples across the disease spectrum and is now accumulating longitudinal samples over repeated annual intervals (NCT02855476).

These insights into the longitudinal dynamics of mHTT and NfL shed light on the biology of HD in human mutation carriers and will be of immediate value in the design and conduct of disease-modifying clinical trials, especially as we enter the era of prevention trials where qualified surrogate endpoints will be fundamental^24^. Looking ahead, some centres are already incorporating blood NfL measurement into shared clinical decision-making in neurological disease^19^. Continued study may reveal a role for mHTT and NfL in guiding decision-making for individuals living with HD.

## Methods

A preprint was submitted to medRxiv on 27^th^ March 2020.

### Study design and participants

HD-CSF was a prospective single-site study with standardised longitudinal collection of CSF, blood and phenotypic data (online protocol: DOI: 10.5522/04/11828448.v1). Eighty participants were recruited. All phenotypic assessment measures were predefined for HD-CSF based on metrics shown to have the largest effect sizes for predicting HD progression ^27^.

This study was performed in accordance with the principles of the Declaration of Helsinki, and the International Conference on Harmonization Good Clinical Practice standards. Ethical approval was obtained from the London Camberwell St Giles Research Ethics Committee (15/LO/1917). Prior to undertaking study procedures, all participants gave informed consent which was obtained by clinical staff.

Manifest HD was defined as UHDRS diagnostic confidence level (DCL) of 4 and *HTT* CAG repeat count ≥ 36. PreHD participants had CAG ≥ 40 and DCL < 4. Controls were age- and gender-matched to mutation carriers, mostly spouses or gene-negative siblings of HD mutation carriers and with no neurological signs or symptoms. Baseline assessments were conducted from February 2016 to February 2017^14^. 24-month (± 3 months) follow-up was conducted from January 2018 to January 2019. At baseline, participants were invited to undergo an optional repeat sampling 4-8 weeks after baseline.

Motor, cognitive and functional status were assessed using the UHDRS from the core Enroll-HD battery, including: the UHDRS Total Motor Score, Total Functional Capacity, Symbol Digit Modalities Test, Stroop Word Reading, Stroop Color Naming and Verbal Fluency – Categorical. These were performed at either the screening or an associated Enroll-HD visit (https://www.enroll-hd.org) within the 2 months prior to screening. We employed a calibrated iteration of the composite UHDRS (cUHDRS)^28^. The cUHDRS was chosen as the primary outcome measure for the analysis of clinical progression as it has favourable signal-to-noise characteristics, encompasses clinical deterioration across multiple relevant domains and has regulatory acceptance as a meaningful measure of HD severity (NCT03761849)^18^.

### Sample collection and processing

Sample collection and processing were as previously described^29^. All collections were standardised for time of day after overnight fasting and processed within 30 minutes of collection using standardised equipment. Blood was collected within 10 minutes of CSF and processed to plasma.

### Analyte Quantification

CSF and plasma NfL were quantified in duplicate using the Neurology 4-plex B assay on the Simoa® HD-1 Analyzer (Quanterix, USA), per manufacturer’s instructions. A 4x dilution for blood samples was performed automatically by the HD-1 Analyser and CSF samples were manually diluted 100x in the sample diluent provided prior to loading onto the machine. The limit of detection (LoD) was 0.105 pg/mL and lower limit of quantification (LLoQ) 0.500 pg/mL. NfL was over the LLoQ in all samples. The intra-assay coefficient of variance (CV) (calculated as the mean of the CVs for each sample’s duplicate measurements) for CSF NfL and plasma NfL was 5.0% and 3.7% respectively. The inter-assay CVs (calculated as the mean of the CVs for analogous spiked positive controls provided by the manufacturer and used in each well plate) for CSF NfL and plasma NfL were 2.7% and 8.4% respectively. We previously quantified NfL in the same baseline samples using an ELISA (NF-Light^®^, UmanDiagnostics, Sweden) in CSF and 1-plex Simoa® kit (NF-Light^®^, Quanterix) in plasma^14^. In both biofluids, agreement between assays was good (Supplementary Fig. 2).

CSF mHTT was quantified in triplicate using the same 2B7-MW1 immunoassay as at baseline (SMC^TM^ Erenna® platform, Merck, Germany)^6^. The LoD was 8fM and LLoQ 25fM. All control samples were below the LoD of the assay except one subject’s baseline re-measured sample. These were imputed as 0 fM for analysis purposes. 27 (21%) samples were below the LLoQ and were included in subsequent analyses. One preHD had CSF mHTT below the LoD in their re-measured baseline sample and was not included in the analyses. The intra-assay CV for CSF mHTT was 14.1%. Haemoglobin contamination was quantified using a commercial ELISA (E88-134, Bethyl Laboratories, USA) by Evotec. Only 1 sample (2.186 μg/mL) had haemoglobin just over the 2 μg/mL recommended threshold^30^.

Assays were run using same-batch reagents, blinded to clinical data.

### MRI Acquisition

The MRI acquisition protocol was identical to that used at baseline^14^. T1-weighted MRI data were acquired on a single 3T Siemens Prisma scanner using a protocol optimized for this study. The parameters were as follows: Images were acquired using a 3D magnetization-prepared 180 degrees radio-frequency pulses and rapid gradient-echo (MPRAGE) sequence with a repetition time (TR)=2000ms and echo time (TE)=2.05ms. The acquisition had an inversion time of 850ms, flip angle of 8 degrees, matrix size 256×240mm. 256 coronal partitions were collected to cover the entire brain with a slice thickness of 1.0 mm. Parallel imaging acceleration (GeneRalized Autocalibrating Partial Parallel Acquisition [GRAPPA], acceleration factor [R]=2) was used and 3D distortion correction was applied to all images.

### MRI Processing

Predefined regions-of-interest for volumetric analysis included the caudate, white matter, grey matter and whole brain. All baseline volumes were re-calculated at follow-up. Bias correction was performed on all scans prior to processing using the N3 procedure^31^. All scans, segmentations and registrations underwent visual quality control blinded to group status to ensure successful processing. All T1-weighted scans passed visual quality control check for the presence of significant motion or other artefacts before processing; one scan failed quality control due to the presence of significant motion, meaning that 57 scans were processed. As described previously, a semi-automated segmentation procedure was performed via Medical Image Display Analysis Software (MIDAS)^32^ to generate volumetric regions of the whole-brain and Total Intracranial Volume (TIV) at baseline^14^. Changes in whole-brain and caudate were calculated via the Boundary Shift Integral (BSI) method^33,34^. The BSI is a semi-automated technique applied within MIDAS that quantifies change over time in regions of interest. For the whole-brain, baseline and follow-up scans were segmented with MIDAS via a morphological segmentor that uses the application of operator-driven thresholds and erosions and dilations to separate brain tissue from the scalp and CSF^32^. The baseline and follow-up scans were then registered using 12 degrees-of-freedom and the BSI metrics were calculated for each participant^35^. One scan failed registration and thus was excluded from the measures of whole-brain change.

Caudate volumes were generated using the automated MALP-EM software^36^. The caudate regions from this procedure underwent visual quality control and were used to calculate the caudate BSI (CBSI) based on a previously validated procedure^34^. This procedure uses local rigid registrations to align the caudate region between baseline and follow-up scans, with a separate registration for left and right caudate. Measures of caudate change are then calculated between the two time points for each participant. No registrations failed quality control.

Baseline grey/white matter volumes were measured via voxel-based morphometry^37^. To calculate grey and white matter change, a fluid-registration approach was applied^38–40^. Baseline and follow-up scans were registered using fluid registration, which ran for 300 iterations^41^. The result of this registration was a voxel compression map (VCM) for each participant, representing the amount of contraction or expansion required within each voxel to map the follow-up scan to the baseline scan. Baseline grey and white matter regions, segmented as described in Byrne et al. (2018)^14^, were convolved with the VCM for each participant to calculate volume change within each tissue class for each participant. Registration failed for three datasets, resulting in the analysis of 55 scan pairs.

Cross-sectional data from the follow-up time point were used to replicate the baseline results. Follow-up whole-brain volume was measured via the semi-automated procedure described at baseline. Follow-up caudate volume was computed by the baseline volume minus the amount of atrophy measured via the CBSI, and follow-up grey and white matter volumes were calculated by subtracting the amount of atrophy from baseline volumes.

### Statistical Analysis

As previously observed^14,15^, NfL distributions were right-skewed, therefore log-transformed values were used for analytical purposes. Due to their known effects on HD, all models included age and CAG repeat count as covariates.

#### Cross-sectional analyses

To validate previous findings and compare assays, we replicated the cross-sectional analyses from the study baseline^14^, using re-measured baseline data and newly collected 24-month follow-up data. To investigate intergroup differences, we applied generalised linear regression models estimated via ordinary least squares, with analyte concentration as the dependent variable, and group membership, and age as independent variables and then with group membership, age and CAG as independent variables. To study associations in HD mutation carriers between the analytes and clinical or imaging measures we used Pearson’s partial correlations adjusted for age and for age and CAG. Bias-corrected and accelerated bootstrapped 95% confidence intervals (95% CI) were calculated for mean differences and correlation coefficients. To understand the discriminatory power of the studied analytes, we produced receiver operating characteristics (ROC) curves for each analyte to differentiate healthy controls from HD mutation carriers, and premanifest from manifest HD and compared areas under the curves (AUC), formally using the method suggested by DeLong and colleagues^42^.

#### Longitudinal modelling

For modelling analyte trajectories over time generalised mixed effect models were performed, estimated via restricted maximum likelihood, with analyte concentration as the dependent variable. Independent models were developed for healthy controls and HD mutation carriers. Only HD mutation carriers were modelled for mHTT. For CSF mHTT in HD mutation carriers, the model had fixed effects for age and CAG, a random intercept per participant and a random slope for age. A similar model was used for healthy controls for NfL in CSF and plasma. HD mutation carriers were modelled with fixed effects for age (second-order) and CAG, and random slopes for age were included for both CSF and plasma NfL.

#### Change-point analysis

We use an offline Bayesian change-point algorithm to estimate the most likely disease time at which a given biomarker changes from a normal to abnormal state. The algorithm was adapted from Zhou et al (2017)^43^ and estimates the marginal likelihood that the data over a segment of time is generated by a given underlying model. We explicitly model changes in both the mean and covariance and use a minimally informative prior. The change-point is then inferred by a change in likelihood of the underlying model Supplementary Fig. 5. As we want to estimate the point of change from normality to abnormality, we use data from all groups (control, preHD, and HD) to fit the model over each time segment.

#### Rates of change simulations

The longitudinal models above were used to estimate rate of change from simulated data. Model parameters, age and CAG distributions, and sample sizes were mimicked from the HD-CSF cohort. Each simulation was repeated 1,000 times and run independently for each analyte for each participant subgroup (i.e. healthy controls, premanifest and manifest HD).

Associations of the analytes’ baseline values, and of their rates of change, with clinical and imaging changes, were assessed using Pearson’s partial correlations adjusted for age, and for age and CAG. Rates of change were computed as the 24-month follow-up value minus the baseline value divided by the follow-up time in years. Bias-corrected and accelerated bootstrapped 95%CI were calculated for correlation coefficients and mean differences. To further explore clinical prognostic value, we divided mutation carriers into nominally “fast” and “slow” progressors at the previously-described cUHDRS minimal clinically important difference for decline (absolute 1.2-point reduction)^28^. Intergroup differences were investigated with generalized linear regression estimated via ordinary least squares, with analyte concentration or rate of change as dependent variable, and group membership and age, and then group, age and CAG as independent variables.

Receiver operating characteristic (ROC) curves were produced and areas under the curves (AUC) compared for the ability of analytes’ baseline values and rates of change to differentiate healthy controls from mutation carriers, and preHD from manifest HD using the method of DeLong^42^.

Clinical trial simulation: We used 1,000 repetitions, a parallel design without attrition or placebo effect, a pseudo-control arm emulating the observed longitudinal trajectories, and an intervention arm with constant 20% annualized reduction in the analyte of interest. Synthetic datasets were generated with Monte Carlo simulations using mixed effect models matching the longitudinal models above. Main effects were estimated as inter-arm mean difference in the mean change from baseline, adjusted for CAG using generalized linear models estimated as above.

Data were analysed using StataMP 16 (StataCorp, USA).

### Event-based modelling

We used an event-based model (EBM)^44^ to estimate the most likely sequence of biomarker changes and to stage participants at both baseline and follow-up. In brief, the EBM is a probabilistic model of observed data generated by an unknown sequence of biomarker events, where an event is defined as a biomarker transitioning from a normal to an abnormal state. The model learns the biomarker distributions of normality and abnormality directly from data, and hence estimates the most likely sequence of abnormality over the whole population. The EBM has been applied extensively to several progressive neurological diseases, including Alzheimer’s disease, multiple sclerosis and HD^45–47^.

We recently developed an EBM for HD biofluid, neuroimaging and clinical biomarkers using baseline data from the HD-CSF cohort^14^. Here we refit the model using baseline data from participants who are present at both baseline and follow-up, and use this model to both test the sequence of events estimated in Byrne et al., (2018)^14^, and to stage participants at both time-points. Specifically, mixture models^47^ were fit to distributions of healthy control and manifest HD participants for each biomarker separately. All biomarkers were adjusted for age, sex, and total intracranial volume in the healthy control cohort at baseline. Following Wijeratne et al. (2018)^47^, the fitted mixture models and a uniform prior on the initial stage were used to estimate the maximum likelihood sequence of events, and its uncertainty estimated using Markov Chain Monte Carlo sampling of the posterior. Participants were then staged by their maximum likelihood position in the baseline sequence.

### Random forest analyses

To complement our analyses, we used random forest methodology, a supervised ensemble - learning approach based on building multiple independent decision trees from bootstrapped samples^48,49^. This allows the detection of non-linear relationships, and simultaneous ranking of predictors of clinical change in multivariate analyses. We implemented random forests on the following outcomes: annualised rate of change in cUHDRS and cUHDRS as a binary outcome: “fast” and “slow” progressors defined as above. Models were first run with biofluid biomarkers including their baseline value and annualised rate of change, and then with biofluid and imaging biomarkers baseline value as predictor variables. Age, CAG repeat count, and DBS were included in all models to serve as comparators variables.

Each random forest had 1,000 trees and was based on bootstrapped samples with replacement and three randomly sampled predictor variables were considered for splitting each node. Relative importance rankings were based on the mean decrease Gini score across all trees, where a higher mean decrease in Gini indicates greater predictor variable relative importance at predicting outcomes. To explicitly test the stability of our results and generate ranking distributions, we re-ran the model 100 times, each containing 80% of the possible observations (randomly selected). Random forests were implemented using R randomForest package^49^.

## Data Availability

The data that support the findings of this study are available on request from the corresponding author, EJW. The data are not publicly available due to their containing information that could compromise the privacy of research participants.

## Acknowledgements

We would like to thank all the participants from the HD community who donated samples and gave their time to take part in this study.

This work was supported by the Medical Research Council UK, the CHDI foundation, the Huntington’s disease Society of America, the Hereditary Disease Foundation, the Wellcome Trust (Wellcome Collaborative Award In Science 200181/Z/15/Z and Wellcome/EPSRC Centre for Medical Engineering [WT 203148/Z/16/Z]), the Department of Health’s NIHR Biomedical Research Centres funding scheme, the UK Dementia Research Institute that receives its funding from DRI Ltd., Alzheimer’s Society, Alzheimer’s Research UK, F. Hoffmann-La Roche Ltd, Horizon 2020 Framework Programme, Engineering and Physical Sciences Research Council, the Swedish Research Council, the European Research Council, and Swedish State Support for Clinical Research, and the Innovative Medicines Initiative Joint Undertaking under EMIF grant.

## Author Contributions

EJW designed the study with the input of HZ, RIS, and SJT. FBR and LMB were involved in participant recruitment. Eligibility, clinical examinations and sample collection were performed by FBR, LBM, and RT. Imaging assessments were conceived RIS, EBJ and EDV, data was acquired by EDV, MA, EBJ, and processed by RIS and EBJ. AH and LMB processed and analysed the patient samples. FBR developed and performed the statistical analysis; PAW developed and performed the EBM and change-point analyses; NG and RH developed and performed the random forest analysis. FBR, LMB and EJW interpreted the data and wrote the manuscript; and all authors contributed to reviewing the manuscript.

## Extended data

**Extended Data Fig. 1.**
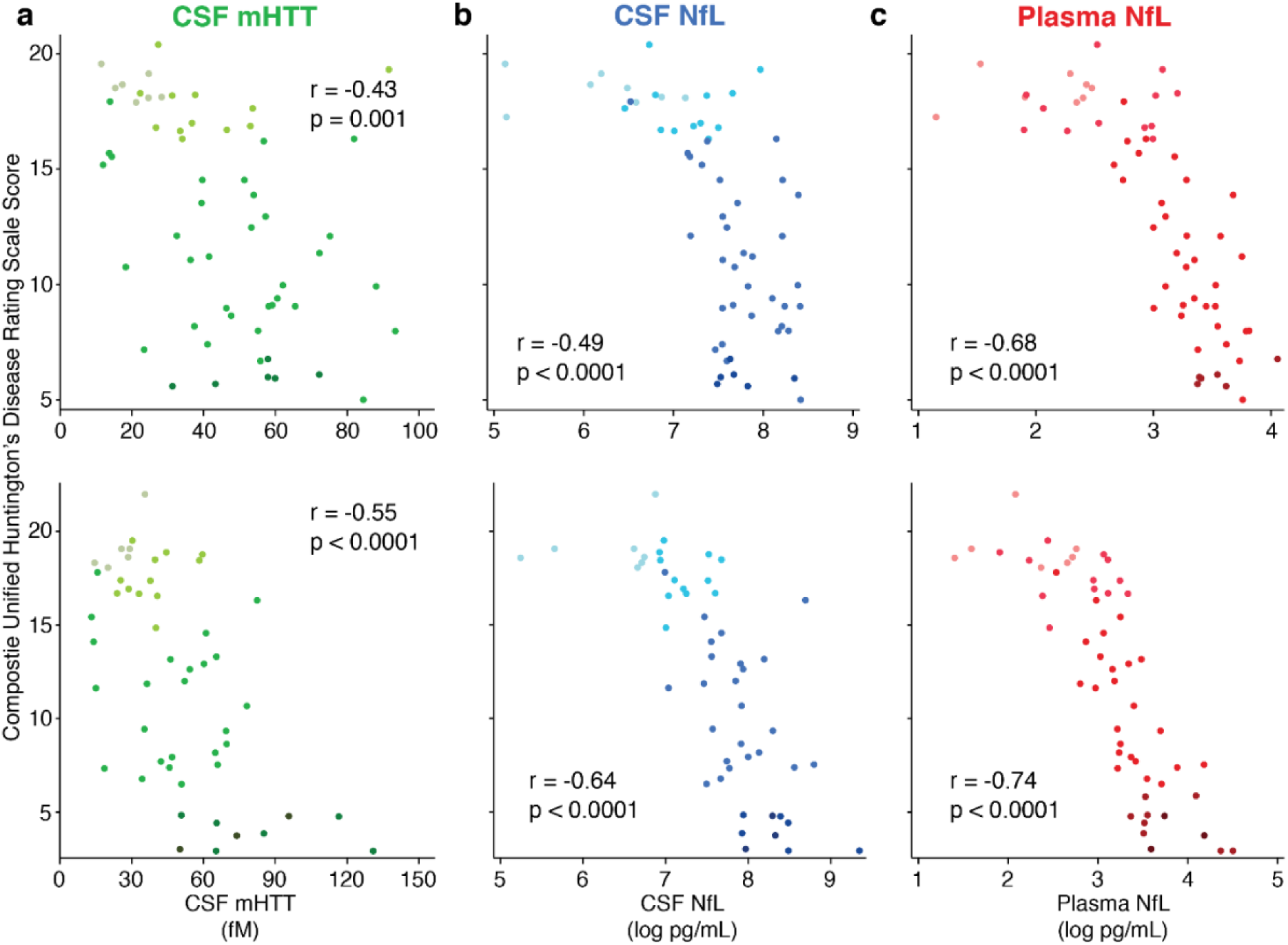
Cross-sectional associations with cUHDRS at baseline and 24-month follow-up. Association within HD mutation carriers between CSF mHTT (green; a, d), CSF NfL (blue; b, e), plasma NfL (red; c, f) and cUHDRS score at baseline (a-c; n=58; n=59; n=59 respectively) and 24-month follow-up (d-f; n=51; n=52; n=54 respectively). Scatter plots show unadjusted values. r and p values are age-adjusted, generated from Pearson’s partial correlations including age as a covariate. NfL values are natural log transformed. UHDRS Unified Huntington’s Disease Rating Scale; PreHD, premanifest Huntington’s disease; HD, Huntington’s disease; CSF, cerebrospinal fluid; mHTT, mutant huntingtin; NfL, neurofilament light.

**Extended Data Fig. 2.**
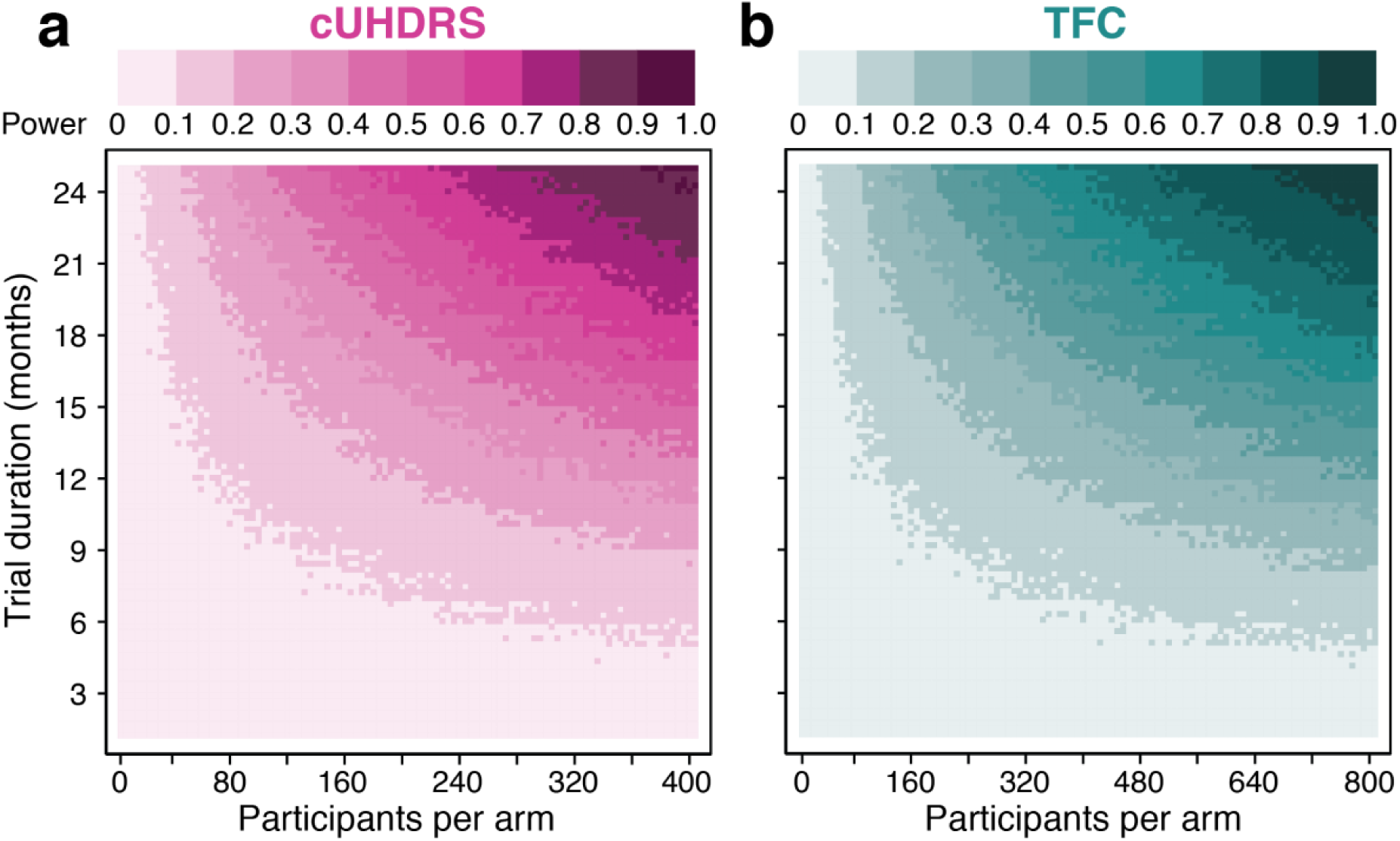
Statistical power, sample size and trial duration of two currently used clinical scales. Monte Carlo simulations predicting the statistical power of using (a) cUHDRS and (b) TFC in the clinical trial context, contingent on sample size per arm and trial duration in months. 2-arm parallel design clinical trials with no attrition or placebo effect with a constant effect size of 20% reduction in each analyte per year were simulated. Each pixel represents 1,000 simulated clinical trials, generated using generalised mixed effects models shaped to estimate the longitudinal trajectories of each clinical scale the main effect in each simulation repetition was calculated as the inter-arm mean difference in the mean change from baseline, using generalised linear models adjusted for CAG. Statistical power was calculated as the proportion of trial simulations with a p-value < 0.05 for the main effect. cUHDRS, Composite Unified Huntington’s disease rating scale; TFC, Total Functional Capacity.

**Extended Data Fig. 3.**
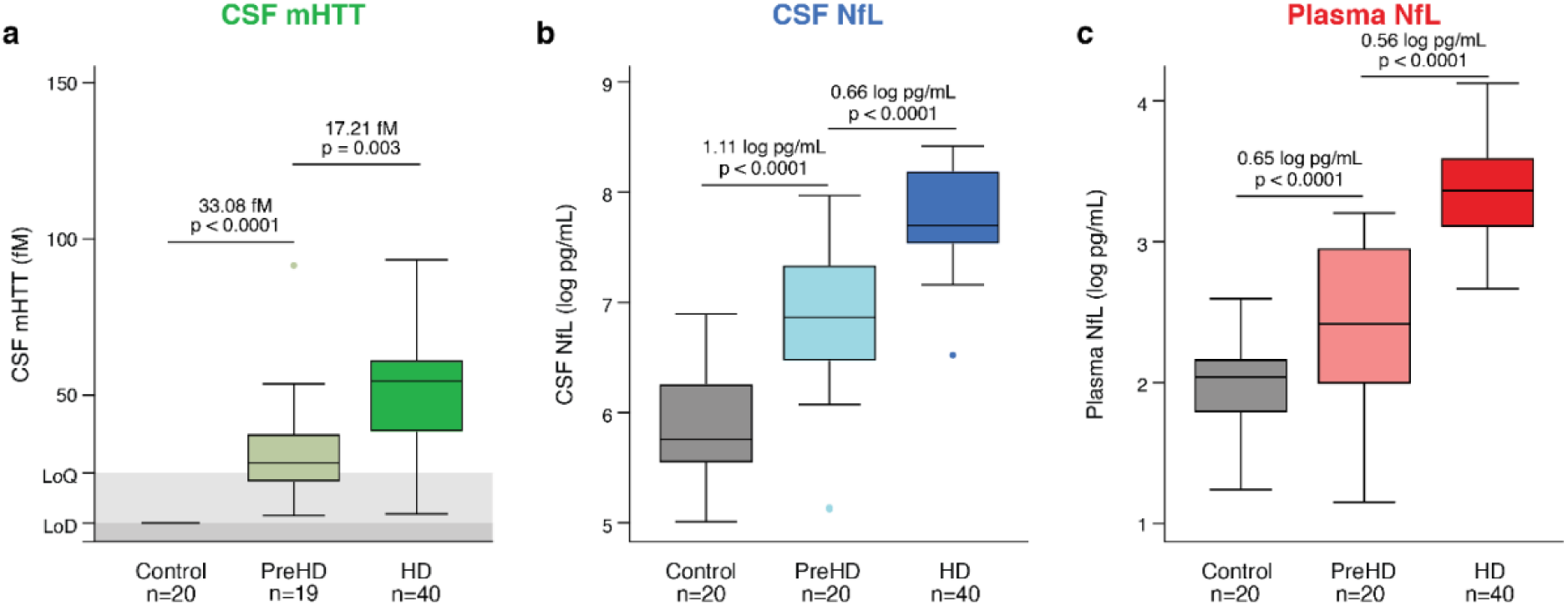
Cross-sectional disease group comparisons in re-measured baseline samples. Concentration of (a) CSF mHTT, (b) CSF NfL, (c) plasma NfL in healthy controls, premanifest HD (PreHD) and manifest HD (HD) participants. NfL values are natural log transformed. Mean differences and p-values were generated from multiple linear regression adjusted for age. PreHD, premanifest Huntington’s disease; HD, Huntington’s disease; CSF, cerebrospinal fluid; mHTT, mutant huntingtin; NfL, neurofilament light.

**Extended Data Fig. 4.**
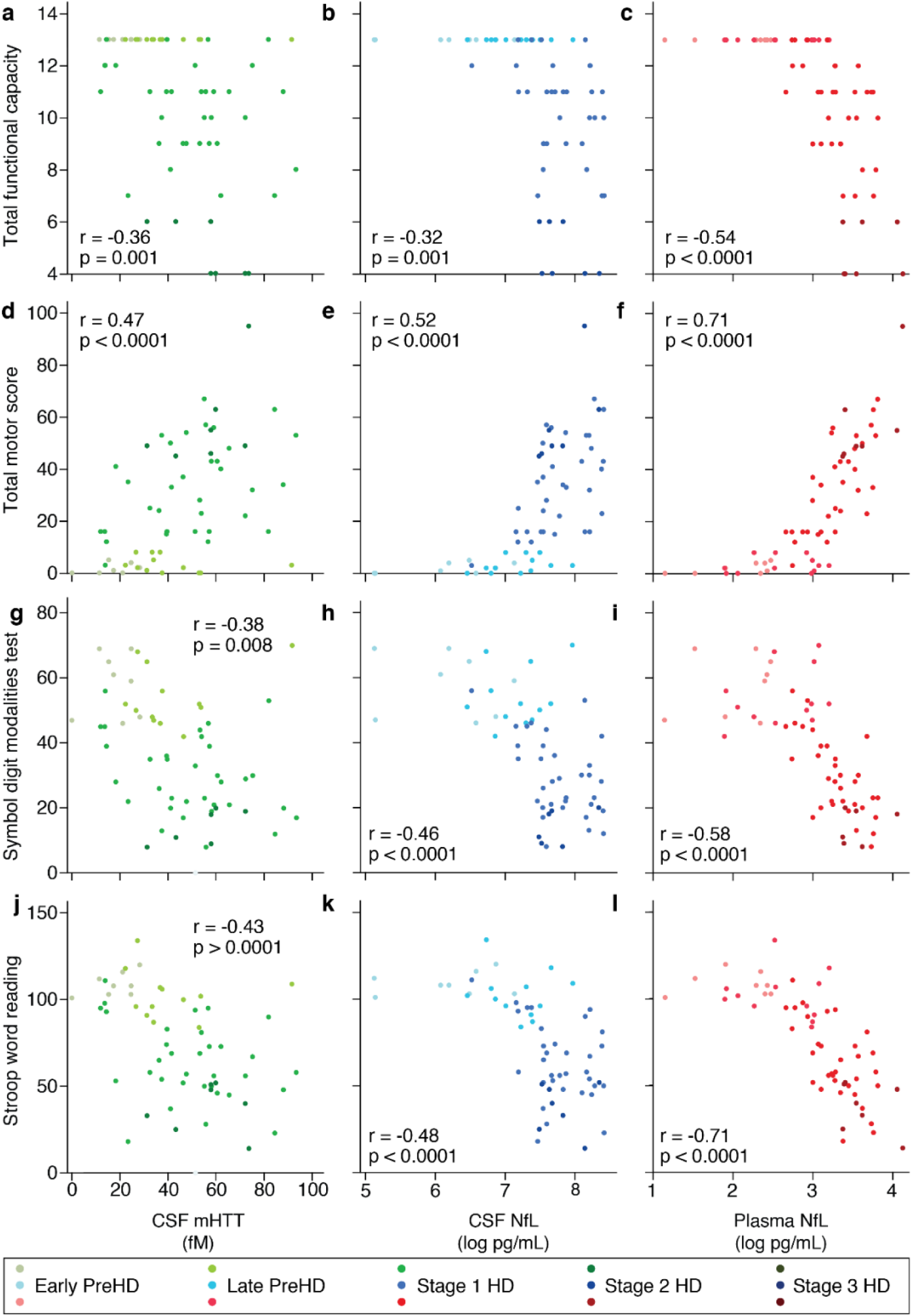
Cross-sectional clinical associations in re-measured baseline samples. Association within HD mutation carriers (n=60) between CSF mHTT (green; a, d, g, j), CSF NfL (blue; b, e, h, k), plasma NfL (red; c, f, i, l) and UHDRS clinical scores including functional (a-c), motor (d-f) and cognitive (g-l) measures. Scatter plots show unadjusted values. r and p values are age-adjusted, generated from Pearson’s partial correlations including age as a covariate. NfL values are natural log transformed. UHDRS Unified Huntington’s Disease Rating Scale; PreHD, premanifest Huntington’s disease; HD, Huntington’s disease; CSF, cerebrospinal fluid; mHTT, mutant huntingtin; NfL, neurofilament light.

**Extended Data Fig. 5.**
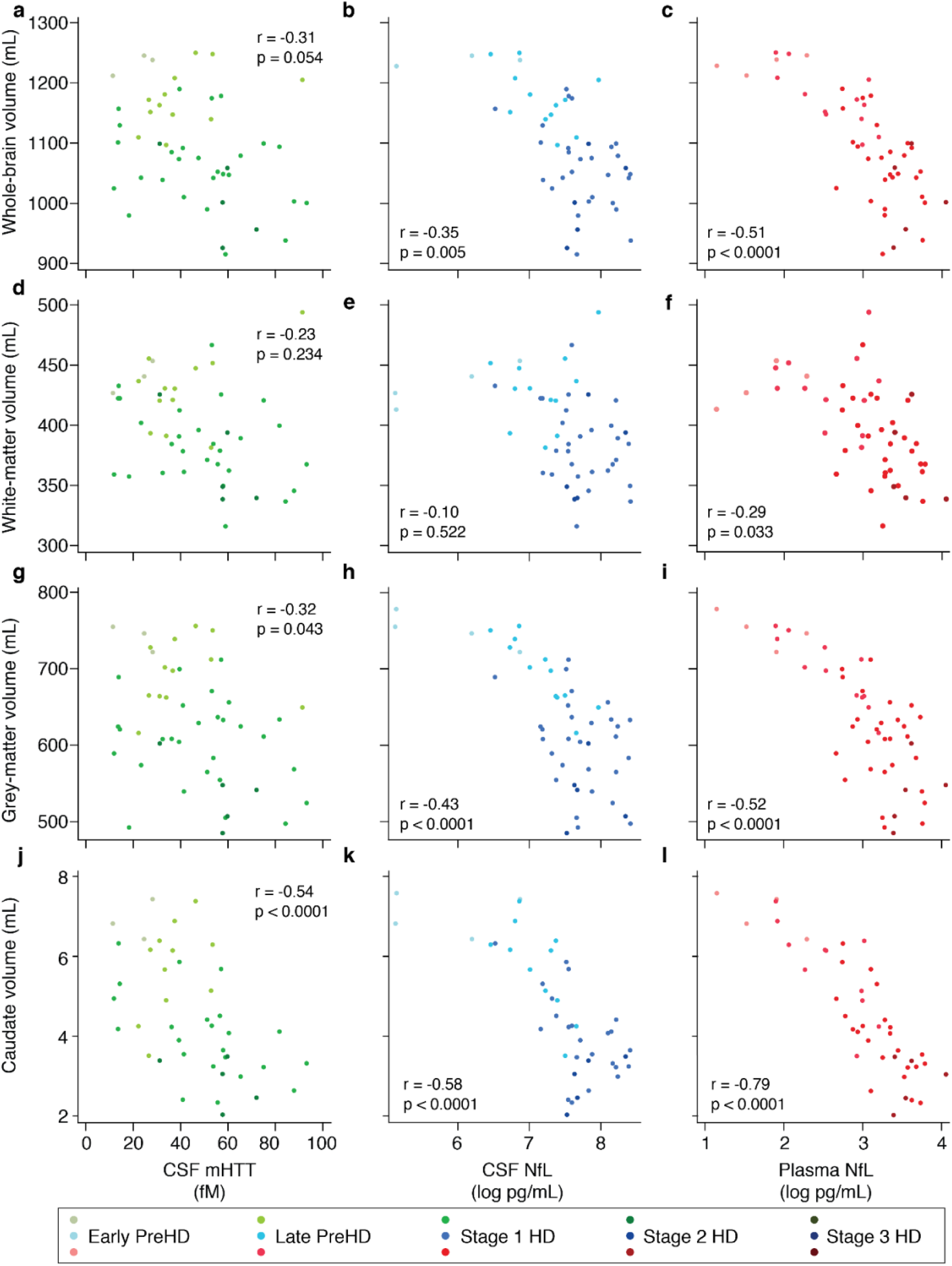
Cross-sectional associations in re-measured baseline samples between analyte concentrations and imaging measures. Association within HD mutation carriers between the analytes CSF mHTT (green; a, d, g, j), CSF NfL (blue; b, e, h, k), plasma NfL (red; c, f, i, l) and MRI volumetric measures whole-brain (n=48; a-c), white-matter (n=49; d-f), grey-matter (n=49; g-i) and caudate (n=43; j-l). All volumetric measures were calculated as a percentage of total intracranial volume. Scatter plots show unadjusted values. r and p values are age-adjusted, generated from Pearson’s partial correlations including age as a covariate. NfL values are natural log transformed. PreHD, premanifest Huntington’s disease; HD, Huntington’s disease; CSF, cerebrospinal fluid; mHTT, mutant huntingtin; NfL, neurofilament light.

**Extended Data Fig. 6.**
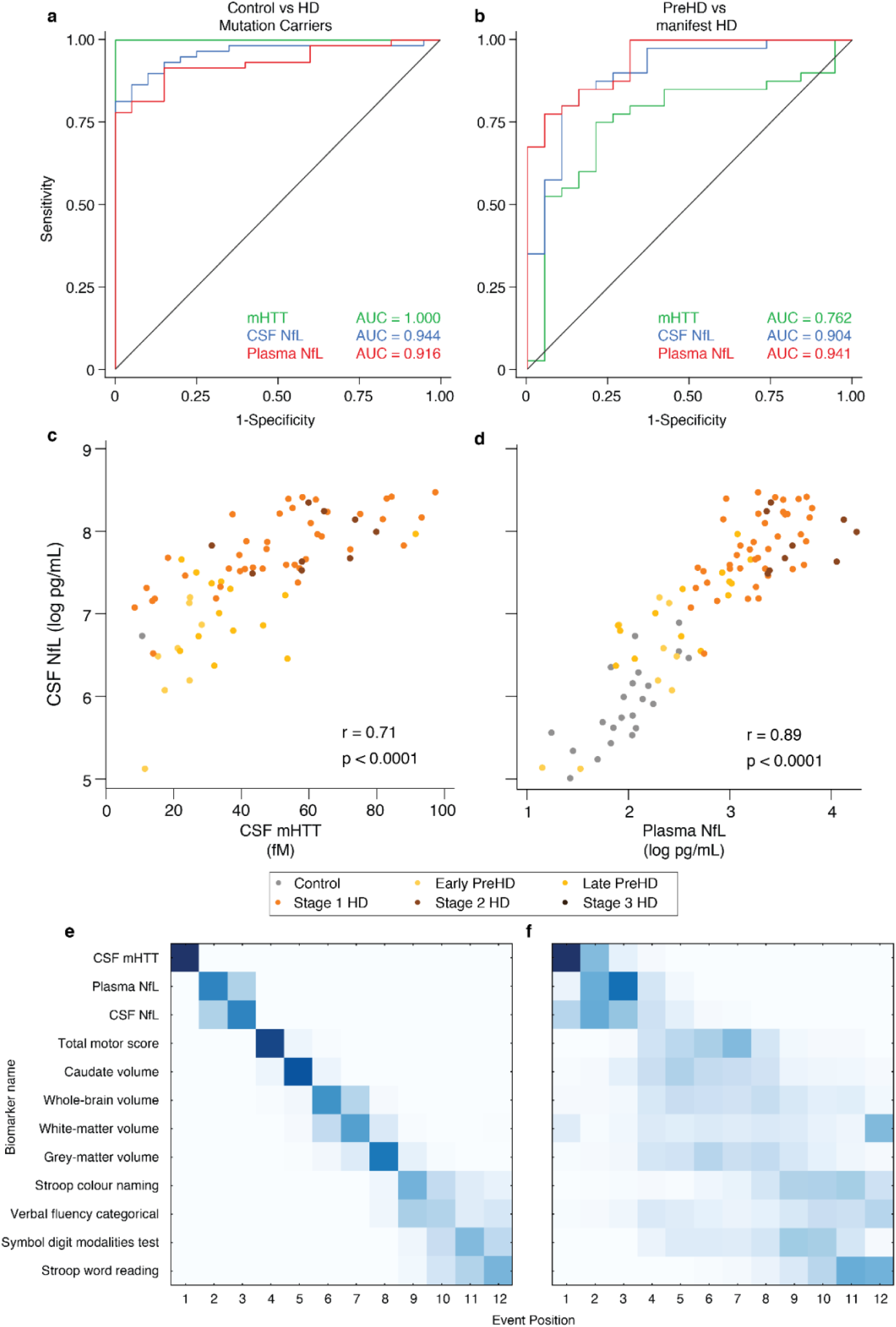
Receiver Operating Characteristics (ROC) analysis, analyte correlations and Event-Based Modelling (EBM) in re-measured baseline samples. ROC curves for the (a) discrimination between controls (N=20) and HD mutation carriers (n=60) (95% CIs for AUCs: CSF mHTT 1.000 – 1.000; CSF NfL 0.894 – 0.994; Plasma NfL 0.855 – 0.977) and (b) discrimination between premanifest (n=20) and manifest HD mutation carriers (n=40) (95% CIs for AUCs: CSF mHTT 0.628 – 0.896; CSF NfL 0.819 – 0.988; Plasma NfL 0.887 – 0.996). Scatter plots showing correlation between CSF mHTT and CSF NfL concentration (c, n=60) and between CSF NfL and Plasma NfL (d, n=80). Scatter plots show unadjusted values. r and p values are unadjusted, generated from Pearson’s correlations. (e) Positional variance diagram produced from the, applied to the 63 HD-CSF participants who had data for all biomarkers (Controls 15; preHD 16; manifest HD 32). (f) Re-estimation of the positional variance in e, using 100 bootstrap samples of the data, providing internal validation of the model’s findings. The positional variance diagrams represent the sequence of “events” (the individual measures going from normal to abnormal, identified by the EBM). Darker diagonal squares represent higher certainty of the biomarker becoming abnormal at the corresponding event where multiple event boxes coloured indicating more uncertainty about its position. 1 indicates the earliest event. NfL values were natural-log transformed. AUC, area under the curve; PreHD, premanifest HD mutation carriers; CSF, cerebrospinal fluid; mHTT, mutant huntingtin; NfL, neurofilament light.

**Extended Data Fig. 7.**
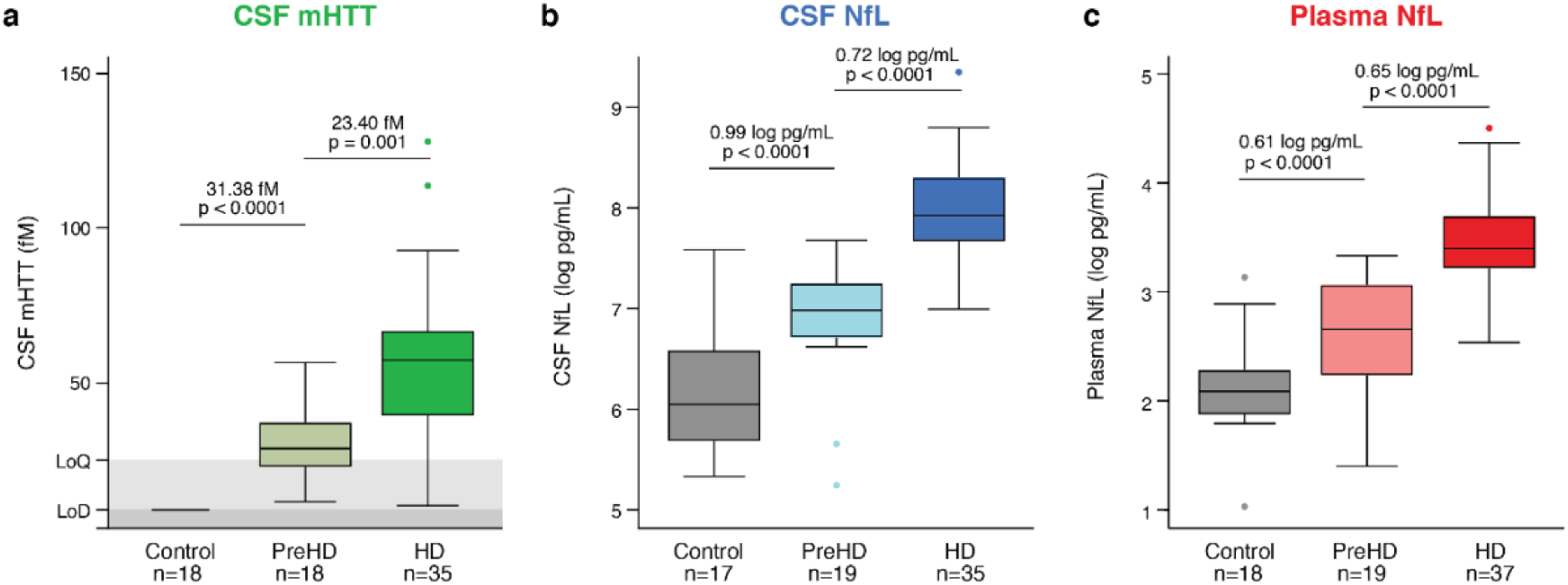
Cross-sectional disease group comparisons in 24-month follow-up samples. Concentration of (a) CSF mHTT, (b) CSF NfL, (c) plasma NfL in healthy controls, premanifest HD (PreHD) and manifest HD (HD) participants. NfL values are natural log transformed. Mean differences and p-values were generated from multiple linear regression adjusted for age. PreHD, premanifest Huntington’s disease; HD, Huntington’s disease; CSF, cerebrospinal fluid; mHTT, mutant huntingtin; NfL, neurofilament light.

**Extended Data Fig. 8.**
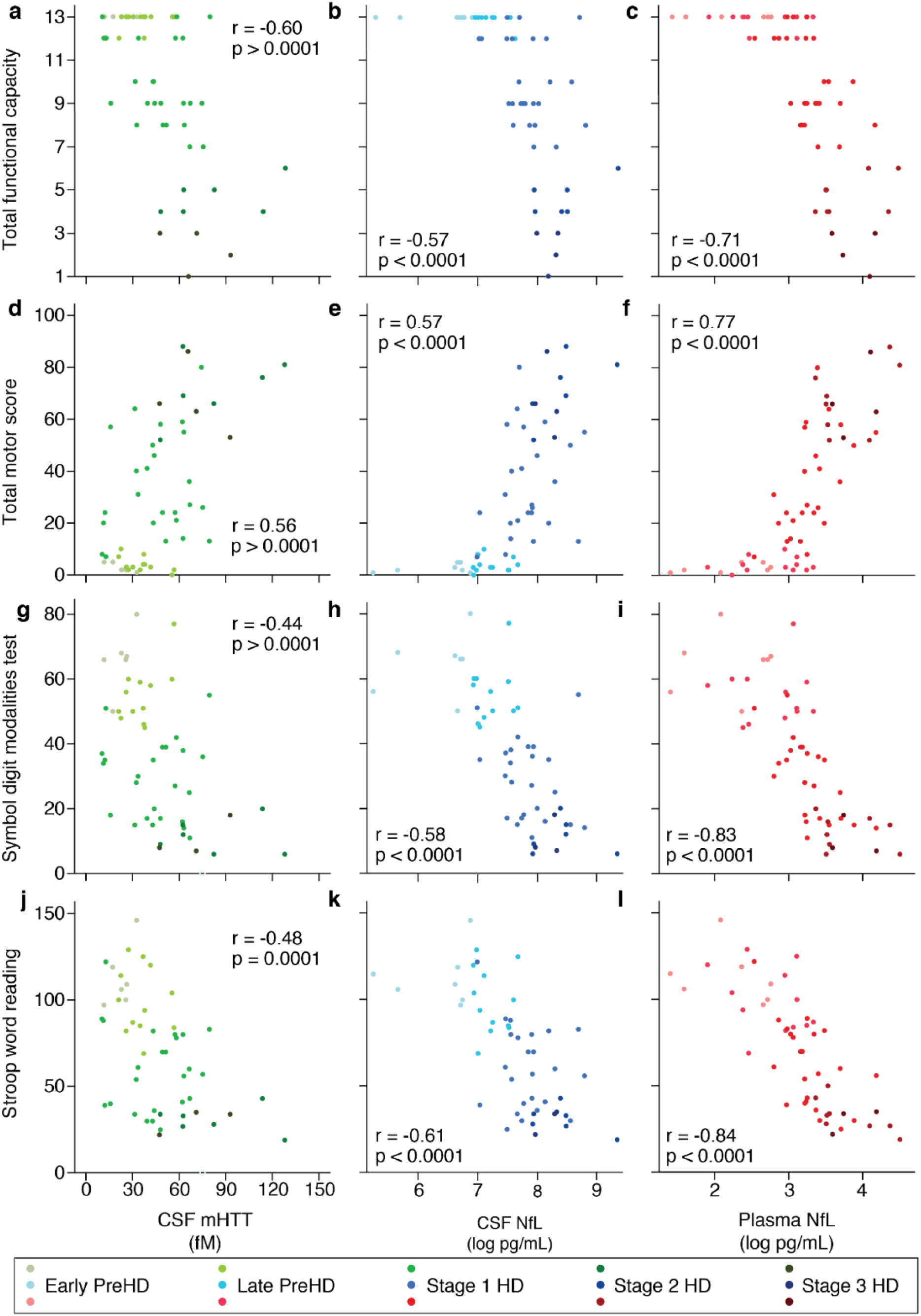
Cross-sectional clinical associations in 24-month follow-up samples. Association within HD mutation carriers (n=60) between CSF mHTT (green; a, d, g, j), CSF NfL (blue; b, e, h, k), plasma NfL (red; c, f, i, l) and UHDRS clinical scores including functional (a-c), motor (d-f) and cognitive (g-l) measures. Scatter plots show unadjusted values. r and p values are age-adjusted, generated from Pearson’s partial correlations including age as a covariate. NfL values are natural log transformed. UHDRS Unified Huntington’s Disease Rating Scale; PreHD, premanifest Huntington’s disease; HD, Huntington’s disease; CSF, cerebrospinal fluid; mHTT, mutant huntingtin; NfL, neurofilament light.

**Extended Data Fig. 9.**
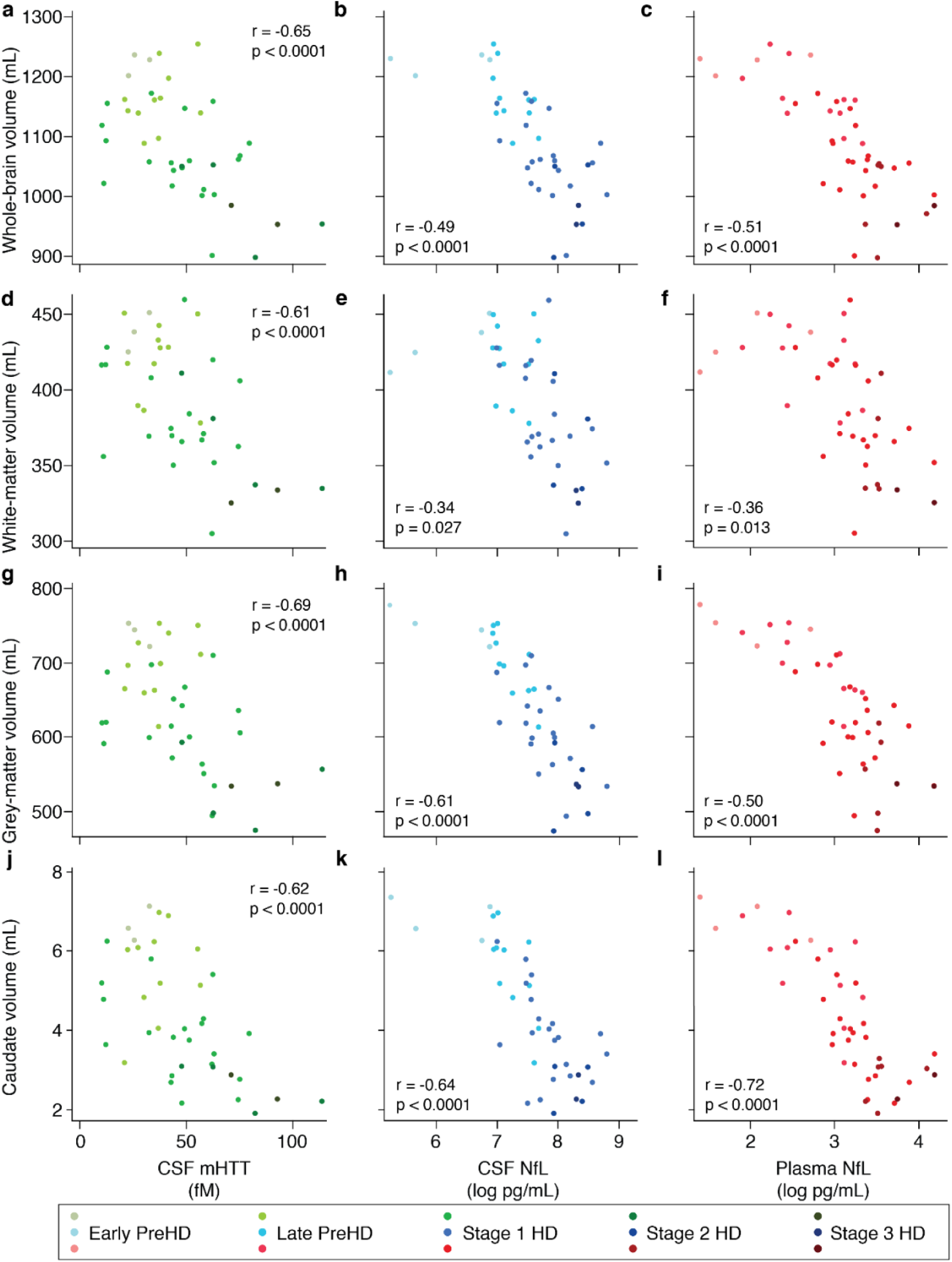
Cross-sectional associations in 24-month follow-up baseline samples between analyte concentrations and imaging measures. Association within HD mutation carriers between the analytes CSF mHTT (green; a, d, g, j), CSF NfL (blue; b, e, h, k), plasma NfL (red; c, f, i, l) and MRI volumetric measures whole-brain (n=43; a-c), white-matter (n=41; d-f), grey-matter (n=41; g-i) and caudate (n=43; j-l). All volumetric measures were calculated as a percentage of total intracranial volume. Scatter plots show unadjusted values. r and p values are age-adjusted, generated from Pearson’s partial correlations including age as a covariate. NfL values are natural log transformed. PreHD, premanifest Huntington’s disease; HD, Huntington’s disease; CSF, cerebrospinal fluid; mHTT, mutant huntingtin; NfL, neurofilament light.

**Extended Data Fig. 10.**
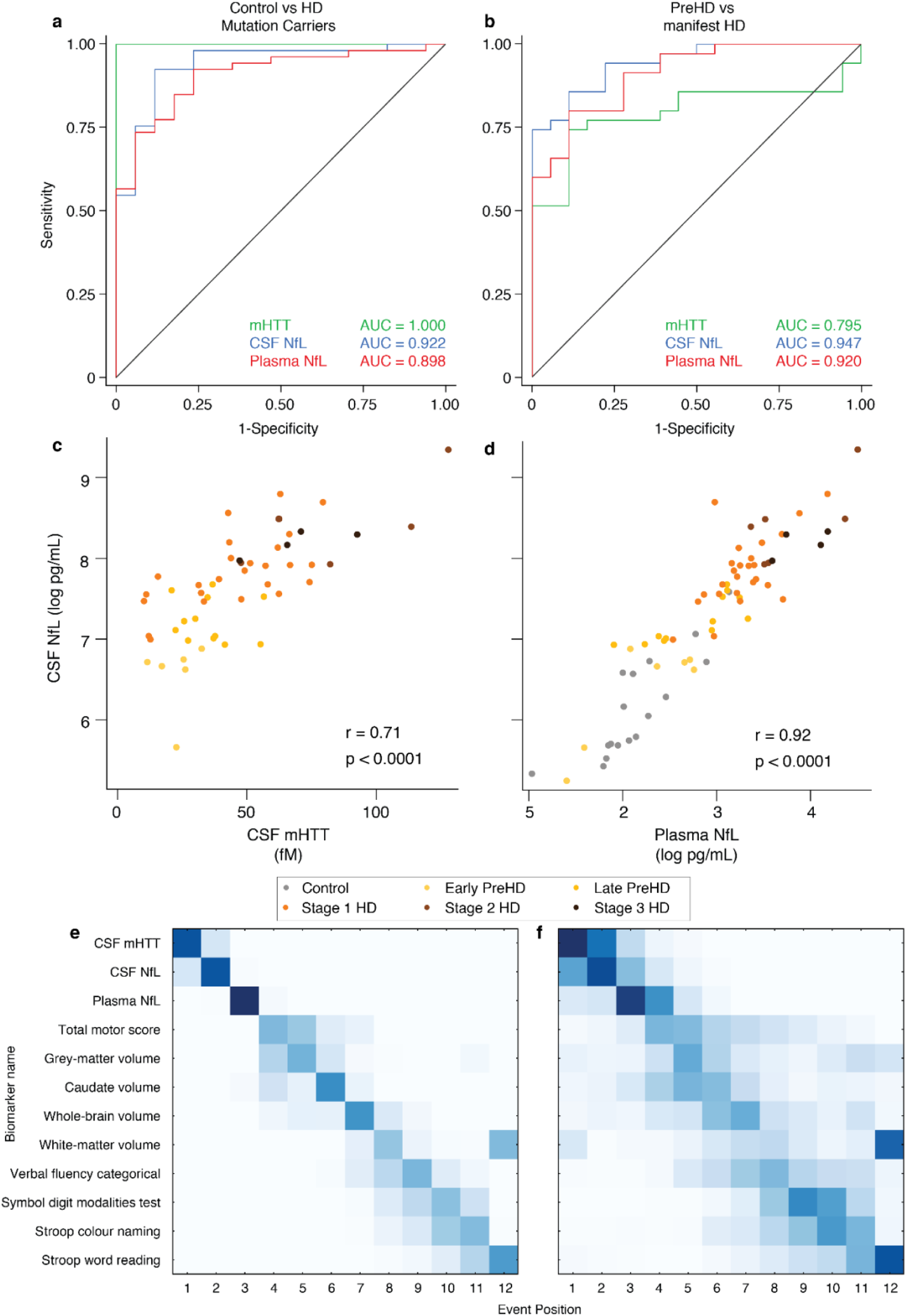
Receiver Operating Characteristics (ROC) analysis, analyte correlations and Event-Based Modelling (EBM) in 24-month follow-up samples. ROC curves for the (a) discrimination between controls (n=17) and HD mutation carriers (n=54) (95% CIs for AUCs: CSF mHTT 1.000 – 1.000; CSF NfL 0.849 – 0.994; Plasma NfL 0.824 – 0.972) and (b) discrimination between premanifest (n=19) and manifest HD mutation carriers (n=35) (95% CIs for AUCs: CSF mHTT 0.672 – 0.919; CSF NfL 0.895 –1.000; Plasma NfL 0.852 – 0.989). Scatter plots showing correlation between CSF mHTT and CSF NfL concentration (c, n=54) and between CSF NfL and Plasma NfL (d, n=71). Scatter plots show unadjusted values. r and p values are unadjusted, generated from Pearson’s correlations. (e) Positional variance diagram produced from the, applied to the 63 HD-CSF participants who had data for all biomarkers (Controls 15; preHD 16; manifest HD 32). (f) Re-estimation of the positional variance in e, using 100 bootstrap samples of the data, providing internal validation of the model’s findings. The positional variance diagrams represent the sequence of “events” (the individual measures going from normal to abnormal, identified by the EBM). Darker diagonal squares represent higher certainty of the biomarker becoming abnormal at the corresponding event where multiple event boxes coloured indicating more uncertainty about its position. 1 indicates the earliest event. NfL values were natural-log transformed. AUC, area under the curve; PreHD, premanifest HD mutation carriers; CSF, cerebrospinal fluid; mHTT, mutant huntingtin; NfL, neurofilament light.

